# The mediating role of epigenetic clocks underlying educational inequalities in mortality: a multi-cohort study

**DOI:** 10.1101/2021.07.01.21259023

**Authors:** Giovanni Fiorito, Sara Pedron, Carolina Ochoa-Rosales, Cathal McCrory, Silvia Polidoro, Yan Zhang, Pierre-Antoine Dugué, Scott Ratliff, Wei Zhao, Gareth J McKay, Giuseppe Costa, Maria Giuliana Solinas, Kathleen Mullan Harris, Rosario Tumino, Sara Grioni, Fulvio Ricceri, Salvatore Panico, Hermann Brenner, Lars Schwettmann, Melanie Waldenberger, Pamela R Matias-Garcia, Annette Peters, Allison Hodge, Graham G Giles, Lauren L. Schmitz, Morgan Levine, Jennifer Smith, Yongmei Liu, Frank Kee, Ian Young, Bernadette McGuinness, Amy Jayne McKnight, Joyce van Meurs, Trudy Voortman, Rose A Kenny, Lifepath consortium, Paolo Vineis, Cristian Carmeli

## Abstract

Educational inequalities in mortality have been observed for decades, however the underlying biological mechanisms are not well known. We assessed the mediating role of altered aging of immune cells functioning captured by DNA methylation changes in blood (known as epigenetic clocks) in educational associated all-cause mortality. Data were from eight prospective population-based cohort studies, representing 13,021 participants. We found educational inequalities in mortality were larger for men than for women, estimated by hazard differences and ratios. Epigenetic clocks explained approximately 50% of educational inequalities in mortality for men, while the proportion was small for women. Most of this mediation was explained by differential effects of unhealthy lifestyles and morbidities of the WHO risk factors for premature mortality. These results support DNA methylation-based epigenetic aging as a signature of educational inequalities in life expectancy emphasizing the need for policies to address the unequal social distribution of these WHO risk factors.

## Introduction

Inequalities in mortality by educational attainment have been observed for decades in developed countries (Mackenbach et al. 2016, Murtin et al. 2017). In some countries these inequalities are widening (Bosworth 2018). The detrimental effect of lower educational attainment on mortality operates through various mediating pathways, including unfavorable working conditions and economic hardship, decreased psychosocial resources, and unhealthy lifestyle (Ross and Wu 1995, Galama, A and van Kippersluis 2018). The concepts of biological embedding or social-to-biological transition describe how social exposures affect various biological processes related to health and survival (Hertzman 2012). Among the proposed biological embeddings, heightened systemic inflammation (Castagné et al. 2016, Berger et al. 2019, Carmeli et al. 2021), dysregulation of physiological systems summarized by allostatic load (Hamdi, South and Krueger 2016, Gruenewald et al. 2012, Castagné et al. 2018), and elevated epigenetic aging of immune cells (Fiorito et al. 2019, Schmitz et al. 2021) have been consistently observed in individuals with lower education in multiple populations. Biomarkers of epigenetic aging from blood-derived DNA methylation profiles have been consistently associated with lifestyle-related behaviors, morbidities, like diabetes, hypertension, and cancer, and are predictors of mortality (Lu et al. 2019, Li et al. 2020, Christiansen et al. 2016, Hillary et al. 2020, Dugué et al. 2018).

Although plausible from previous observations, the mediating role of epigenetic aging in explaining educational inequalities in mortality has been rarely investigated. One study, of older women in the United States of America, found that only a small portion of educational inequalities in mortality was explained by epigenetic aging measured through the DNAmPhenoAge biomarker (Liu et al. 2019). However, the path-specific contribution of epigenetic ageing independent of unhealthy lifestyles and morbidities has not been investigated, thus the independent mediating role of epigenetic aging in social inequality-associated mortality has not been studied.

In this study, we assessed the mediating role of blood DNA methylation-based epigenetic aging between education and all-cause mortality in multiple populations and by sex, as previous observational studies have shown sex differences in educational inequalities in mortality (Galama et al. 2018, Laine et al. 2019). Data spanning the social, biological, and survival layers was derived from eight population-based cohort studies across seven countries, totalling 13,021 participants, followed on average for 9 years from a baseline age of 64. We measured epigenetic age from blood DNA methylation, known as epigenetic clocks. We considered four epigenetic clock biomarkers, namely Horvath’s and Hannum’s DNAmAge (Hannum et al. 2013, Horvath 2013), Levine’s DNAmPhenoAge and Lu’s DNAmGrimAge (Levine et al. 2018, Lu et al. 2019), as they have been commonly studied in the literature, and are associated with education and all-cause mortality in several populations, capturing different aspects of the epigenetic aging processes (Fiorito et al. 2019, McCartney et al. 2020, Lu et al. 2019, Wang et al. 2021). Whether an individual experienced elevated or mitigated epigenetic aging was estimated by computing the residuals from a regression of the epigenetic clock on chronological age. Thereafter, we refer to these values as Horvath DNAmAA, Hannum DNAmAA, DNAmPhenoAA and DNAmGrimAA, respectively. We calculated the portion of educational inequalities in all-cause mortality explained by epigenetic aging biomarkers via a counterfactual-based mediation method (Tchetgen Tchetgen 2013). The chosen method offered the opportunity to incorporate potential exposure-mediator interactions and multiple mediators, and a sensitivity analysis to assess the impact of potentially unmeasured confounding (Nguyen et al. 2015).

Secondly, in this study we assessed the potential path-specific contribution of epigenetic aging independent of unhealthy lifestyle factors (current smoking, low physical activity, high alcohol intake, and high body mass index) and related morbidities (type 2 diabetes and hypertension). We considered these risk factors as additional mediators as they are on the causal path from educational attainment to all-cause mortality (Ross and Wu 1995, Laine et al. 2019, Janke et al. 2020, Brunello et al. 2016) and are targets of policies to reduce premature mortality burden (Stringhini et al. 2017). To address this aim, we considered the joint mediation of these lifestyle-related factors, without the need to specify a causal order among them (VanderWeele and Vansteelandt 2014, Laine et al. 2019).

## Results

### Characteristics of the populations

Summary statistics for each cohort study are presented separately for men and women in **Table 1**. There were more men in MCCS (61%) and more women in EPIC-IT (60%), otherwise there was an equal proportion of the sexes. The average age at baseline ranged from 53 years in EPIC-IT to 70 years in MESA. In all cohorts except MESA, TILDA, and RS (in men) there were more individuals with lower educational attainment than higher. The average follow-up time ranged from 5 years in NICOLA to 15 years in EPIC-IT. Mortality rates were higher for men than women, except in EPIC-IT where they were similar. For women, they ranged from 52 deaths per 10,000 person-years in NICOLA to 262 in ESTHER, while for men mortality rates ranged from 100 in NICOLA to 400 in ESTHER.

**Table 1.** Characteristics of participants (N=13,021). Values for men and women are reported on top and bottom rows, respectively. Mean (standard deviation, SD) age at baseline and length of follow-up are reported in years. Mortality rates per 10,000 person-years are standardized using total person-years by 5-year strata of age at baseline. Characteristics are for participants with complete data on education and mortality.

| Study | Participants<br>N (%) | Age at baseline<br>Mean (SD) | Higher/Lower education<br>N (%) | Death<br>N (%) | Follow-up years<br>Mean (SD) | Mortality rate |
| --- | --- | --- | --- | --- | --- | --- |
| **EPIC-IT** |  |  |  |  |  |  |
| Men | 607 (40) | 53.7 (6.7) | 231 (38) / 376 (62) | 135 (22) | 14.9 (4.4) | 151.2 |
| Women | 917 (60) | 52.9 (7.6) | 255 (28) / 662 (72) | 147 (16) | 15.3 (3.5) | 160.6 |
| **ESTHER** |  |  |  |  |  |  |
| Men | 870 (48) | 62.7 (6.6) | 243 (28) / 627 (72) | 410 (47) | 12.4 (4.9) | 400.5 |
| Women | 944 (52) | 62.2 (6.7) | 210 (22) / 734 (78) | 318 (34) | 13.8 (4.1) | 263.3 |
| **KORA** |  |  |  |  |  |  |
| Men | 840 (49) | 61.3 (8.9) | 373 (45) / 464 (55) | 90 (11) | 8.4 (1.7) | 119.6 |
| Women | 882 (51) | 60.7 (8.8) | 357 (40) / 525 (60) | 56 (6) | 8.7 (1.2) | 89.3 |
| **MCCS** |  |  |  |  |  |  |
| Men | 1719 (61) | 69.2 (7.8) | 619 (36) / 1100 (64) | 756 (44) | 12.6 (5.9) | 248.9 |
| Women | 1093 (39) | 67.7 (8.7) | 314 (29) / 779 (71) | 333 (30) | 13.9 (5.6) | 171.6 |
| **MESA** |  |  |  |  |  |  |
| Men | 612 (49) | 69.7 (9.3) | 528 (86) / 84 (14) | 79 (13) | 6.4 (1.4) | 95.6 |
| Women | 648 (51) | 69.5 (9.4) | 546 (84) / 102 (16) | 58 (9) | 6.5 (1.2) | 65.7 |
| **NICOLA** |  |  |  |  |  |  |
| Men | 963 (49) | 65.7 (9.4) | 374 (39) / 589 (61) | 65 (7) | 4.9 (0.9) | 99.8 |
| Women | 1017 (51) | 63.5 (9.3) | 461 (43) / 556 (57) | 31 (3) | 5.1 (0.7) | 52.0 |
| **RS** |  |  |  |  |  |  |
| Men | 622 (44) | 63.6 (8.1) | 418 (67) / 205 (33) | 79 (13) | 5.9 (2.2) | 231.5 |
| Women | 798 (56) | 64.0 (8.0) | 314 (39) / 484 (61) | 48 (6) | 5.8 (2.0) | 100.9 |
| **TILDA** |  |  |  |  |  |  |
| Men | 244 (50) | 61.9 (8.1) | 148 (61) / 96 (39) | 24 (10) | 7.2 (2.0) | 145.3 |
| Women | 245 (50) | 62.1 (8.4) | 141 (58) / 104 (42) | 11 (5) | 7.2 (1.9) | 68.2 |

### Educational inequalities in all-cause mortality explained by epigenetic aging biomarkers

Men with a lower educational attainment (potentially counter to fact) had an excess mortality from all causes of 55 deaths per 10,000 person-years [95% compatibility interval (CI): 38 to 72] compared with having (potentially counter to fact) higher educational attainment (**Figure 1**). Women with a lower educational attainment (potentially counter to fact) had an excess mortality from all causes of 7 deaths per 10,000 person-years [95% CI: -5 to 20] compared with having higher educational attainment (potentially counter to fact). Compared to the mortality rates for women across the populations (see **Table 1**), this inequality was small. On the relative scale, total effects corresponded to hazard ratios of 1.27 [95% CI: 1.11 to 1.44] and 1.18 [95% CI: 1.01 to 1.37], for men and women respectively (**Figure 2**). Heterogeneity across cohort-specific estimates was low (forest plots in **Supplementary Material)**.

**Figure 1.**
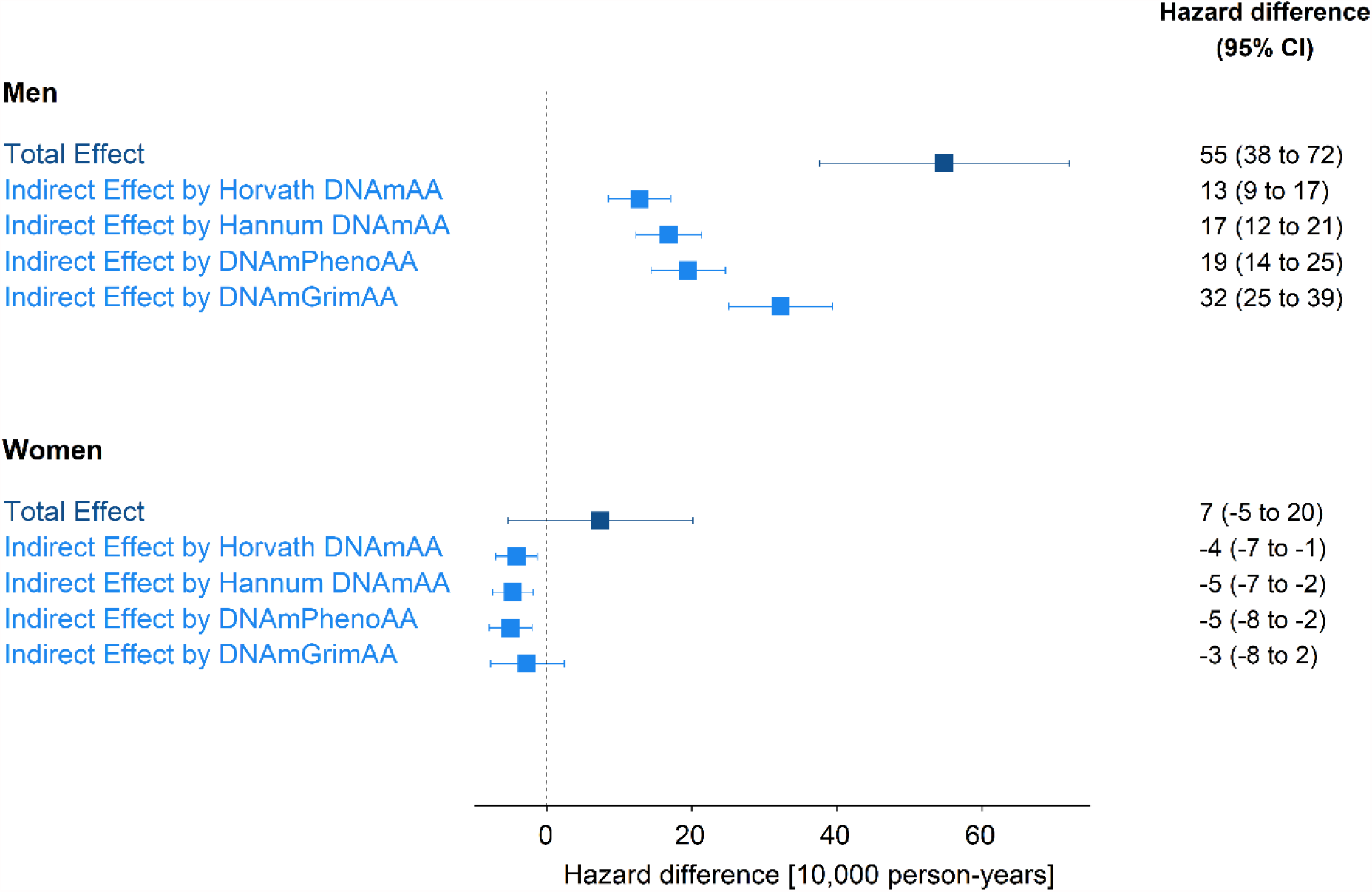
Educational inequalities and mediation by epigenetic aging biomarkers on the absolute scale. Hazard difference per 10,000 person-years and 95% compatibility intervals (CI) are for the total effect of lower (vs higher) educational attainment on all-cause mortality, and for indirect effect by epigenetic aging biomarkers (Horvath DNAmAA, Hannum DNAmAA, DNAmPhenoAA, and DNAmGrimAA). All effects are pooled estimates of single cohort’s hazard differences through a weighted inverse variance meta-analytic model. The total number of participants/deaths across cohorts is 6,477 / 1,638 for men and 6,544 / 1,002 for women.

**Figure 2.**
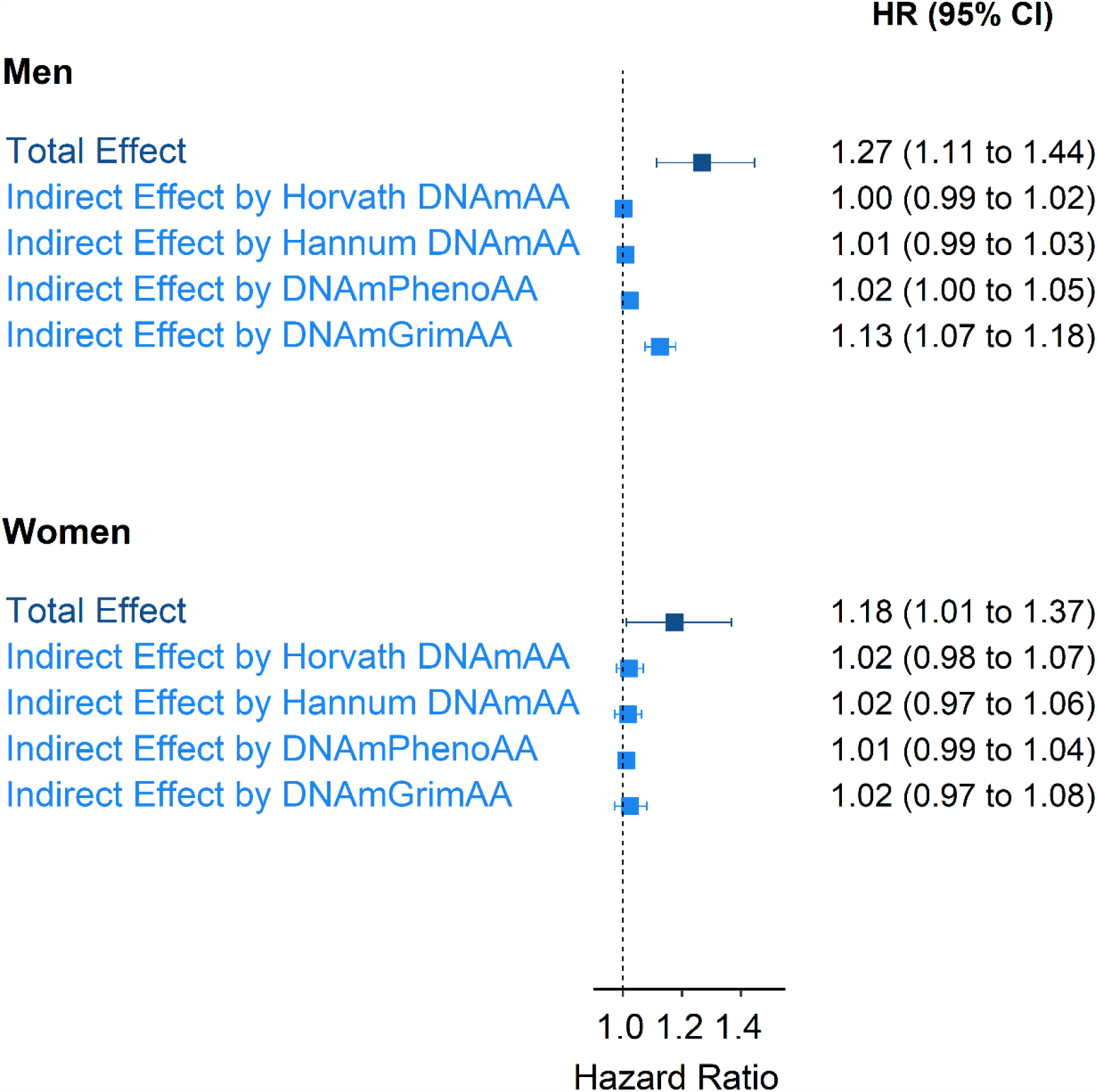
Educational inequalities and mediation by epigenetic aging biomarkers on the relative scale. Hazard ratios (HRs) and 95% compatibility intervals (CI) are for the total effect of lower (vs higher) educational attainment on all-cause mortality, and for indirect effects by epigenetic aging biomarkers (Horvath DNAmAA, Hannum DNAmAA, DNAmPhenoAA, and DNAmGrimAA). All effects are pooled estimates of single cohort’s hazard ratios through a weighted inverse variance meta-analytic model. The total number of participants/deaths across cohorts is 6,477 / 1,638 for men and 6,544 / 1,002 for women.

For men, the portion of the inequalities (total effect described above) explained by epigenetic aging (indirect effect) was equivalent to 13 excess deaths per 10,000 person-years [95% CI: 9 to 17] for Horvath DNAmAA, 17 [95% CI: 12 to 21] for Hannum DNAmAA, 19 [95% CI: 14 to 25] for DNAmPhenoAA, and 32 [95% CI: 25 to 39] for DNAmGrimAA (**Figure 1**). The latter corresponded to 58% of the educational inequalities in mortality (ratio between indirect and total effect). Hazard ratios for indirect effects were small for all but DNAmGrimAA, which corresponded to 1.13 [95% CI: 1.07 to 1.18] (**Figure 2**). This corresponded to 51% of the educational inequalities in mortality on the relative scale (ratio between the logarithm of indirect and total effect).

For women, the portion of the inequalities explained by epigenetic aging biomarkers corresponded to small negative hazard differences and small positive hazard ratios (**Figures 1** and **2**).

### Educational inequalities in all-cause mortality explained by unhealthy lifestyle, morbidities, and epigenetic aging biomarkers

For men, the joint indirect effect explained by unhealthy lifestyle, morbidities and epigenetic aging biomarkers *en-bloc* was equivalent to 29 excess deaths per 10,000 person-years [95% CI: 16 to 41] for Horvath DNAmAA, 31 [95% CI: 20 to 42] for Hannum DNAmAA, 33 [95% CI: 20 to 45] for DNAmPhenoAA, and 36 [95% CI: 24 to 49] for DNAmGrimAA (**Table 2**). On the relative scale, the joint indirect effect resulted in hazard ratios of 1.12 [95% CI: 1.06 to 1.18] for Horvath DNAmAA, 1.12 [95% CI: 1.07 to 1.18] for Hannum DNAmAA, 1.14 [95% CI: 1.07 to 1.18] for DNAmPhenoAA, and 1.18 [95% CI: 1.09 to 1.27] for DNAmGrimAA (**Table 2**). Following the model in **Figure 1B** and the sequential mediation approach described in **Methods**, the estimated path-specific indirect effect by epigenetic aging but not unhealthy lifestyle and morbidities was small for all epigenetic aging biomarkers (**Table 2**), with DNAmGrimAA having the largest path-specific effect of 7.2 excess deaths [95% CI: -4.3 to 18.6] and a hazard ratio of 1.05 [95% CI: 0.98 to 1.13].

**Table 2.** Mediation by unhealthy lifestyle, morbidities, and epigenetic aging in men. Absolute (hazard difference per 10,000 person-years) and relative (hazard ratio) size of total effect of lower (vs higher) educational attainment on all-cause mortality, and of indirect effects by unhealthy lifestyle and morbidities (LM) and epigenetic aging biomarkers (Horvath DNAmAA, Hannum DNAmAA, DNAmPhenoAA, and DNAmGrimAA). All effects are pooled estimates of single cohort’s effects through a weighted inverse variance meta-analytic model. The total number of participants/deaths across cohorts is 6,156 / 1,586.

| Effects | Hazard difference<br>(95% CI) | Hazard ratio<br>(95% CI) |
| --- | --- | --- |
| Total effect | 52 (34 to 69) | 1.27 (1.11 to 1.46) |
| Indirect effect by LM | 29 (18 to 40) | 1.12 (1.06 to 1.17) |
| Indirect effect by LM + Horvath DNAmAA | 29 (16 to 41) | 1.12 (1.06 to 1.18) |
| Indirect effect by Horvath DNAmAA but not LM | -0.8 (-11.8 to 10.2) | 1.00 (0.93 to 1.07) |
| Indirect effect by LM + Hannum DNAmAA | 31 (20 to 42) | 1.12 (1.07 to 1.18) |
| Indirect effect by Hannum DNAmAA but not LM | 1.8 (-9.3 to 12.8) | 1.02 (0.95 to 1.09) |
| Indirect effect by LM + DNAmPhenoAA | 33 (20 to 45) | 1.14 (1.07 to 1.21) |
| Indirect effect by DNAmPhenoAA but not LM | 3.6 (-7.6 to 14.8) | 1.02 (0.95 to 1.09) |
| Indirect effect by LM + DNAmGrimAA | 36 (24 to 49) | 1.18 (1.09 to 1.27) |
| Indirect effect by DNAmGrimAA but not LM | 7.2 (-4.3 to 18.6) | 1.05 (0.98 to 1.13) |

For women, the portion of the inequalities explained by unhealthy lifestyle, morbidities and epigenetic aging biomarkers were small (see **Supplementary Table 3**), with the largest hazard ratio of 1.03 [95% CI: 0.95 to 1.12] for DNAmGrimAA. For this reason, we did not run the sequential mediation analysis.

### Sensitivity analyses

Total and indirect effect estimates were similar when including childhood socioeconomic conditions as a potential confounder of education-mortality, education-epigenetic aging, and epigenetic aging-mortality relationships (**Supplementary Table 4**). When assessing the potential bias due to unmeasured confounding (possibly exposure-induced) of the mediator-outcome association, the indirect effect accounted for by epigenetic aging biomarkers varied by small amounts around the estimates reported in main analysis (**Supplementary Figure 1** for DNAmGrimAA, other biomarkers not shown). This suggested modest bias from unmeasured confounding on the estimated indirect effect by epigenetic aging biomarkers.

By incorporating a non-differential measurement error of three years in epigenetic aging biomarkers, the estimated indirect effects for all epigenetic aging biomarkers were similar to, and for DNAmGrimAA slightly larger than, those reported from main analyses (**Supplementary Figures 2 and 3**). This suggested no overestimation of indirect effects reported from main analyses due to potential measurement error of epigenetic aging.

When assessing potential age-dependent hazard differences/ratios by repeating the analyses with survival ages right censored at 70, 75, 80, and 85 years, educational inequalities in mortality for men on the absolute scale widened (**Supplementary Figure 4**) while they reduced on the relative scale (**Supplementary Figure 5**), particularly between 70 to 80 years. The indirect effect explained by DNAmGrimAA was substantive at all examined ages. For the other biomarkers, the indirect effect was small by 70 years, and slightly increased at later ages. The total and indirect effects for women were small at all ages on the hazard difference scale, while on the hazard ratio scale they were small by 75 years and increased afterwards (especially for the total effect and the indirect effect by DNAmGrimAA). Overall, this was consistent with the findings from the main analyses that educational inequalities in mortality were larger for men than for women on both scales, that DNAmGrimAA explained a substantive portion of those inequalities for men and a small portion on the relative scale for women.

In the sequential mediation analysis to assess the path-specific effect not mediated by leukocyte composition in blood (**Supplementary Figure 6**), for men the indirect effect explained by leukocyte composition in blood was 16 excess deaths per 10,000 person-years [95% CI: 10 to 22], increasing to 34 [95% CI: 26 to 43] when both leukocyte composition in blood and DNAmGrimAA were the mediators (**Supplementary Table 5**). The path-specific indirect effect by DNAmGrimAA but not leukocyte composition was quantified as 18 deaths per 10,000 person-years [95% CI: 10 to 25]. For epigenetic aging based on the other biomarkers, this increment was small. This suggested that the epigenetic aging biomarker DNAmGrimAA captured the differential effect of DNA methylation levels beyond leukocyte composition in blood.

## Discussion

We quantified the contribution of blood DNA methylation-based epigenetic aging biomarkers in explaining educational inequalities in all-cause mortality using data from eight population-based cohort studies across seven high-income countries with a total of 13,021 participants. We found educational inequalities in mortality were larger for men than for women. For men, epigenetic aging as measured by the DNAmGrimAA biomarker explained a substantive portion of educational inequalities in all-cause mortality on both the hazard difference and ratio scales. The other biomarkers explained a smaller portion and were potentially confounded by leukocyte composition in blood. Additionally, we found that this mediation by epigenetic aging biomarkers was mostly explained by differential effects of unhealthy lifestyle (current smoking, high alcohol intake and body mass index, low physical activity) and morbidities (hypertension and diabetes). For women, educational inequalities in all-cause mortality were small on the hazard difference scale and mediation by epigenetic aging biomarkers were small on the hazard ratio scale.

This is the first study to examine the mediating role of biomarkers of epigenetic aging in leukocytes underlying educational inequalities in mortality, by integrating social and lifestyle exposures, DNA methylation profiling from blood, and survival follow-up in men and women from several populations across Europe, USA and Australia. The observed substantive educational inequalities in all-cause mortality for men, whilst smaller for women, were in line with previous reports (Laine et al. 2019, Galama et al. 2018). Among the possible explanations for sex differences are that men with less education are more likely to engage in unhealthy lifestyle behaviors than less educated women (Ross, Masters and Hummer 2012), or that men with less education are more likely to be unemployed than their female counterparts (van Hedel et al. 2018). Additionally, in a sensitivity analysis we found that educational inequalities in mortality widened as a function of age, in line with a previous large European study of 371,295 participants (Gallo et al. 2012).

For men, the portion of educational inequalities in all-cause mortality explained by epigenetic aging was substantive, particularly for the biomarker DNAmGrimAA, on both the absolute and relative scales, and across survival ages from 70 years onwards. Additionally, in sensitivity analysis we found that only DNAmGrimAA captured changes in DNA methylation patterns independently of changes in blood cell composition when mediating the effect of education on mortality. Our finding is consistent with a recent Mendelian Randomization (MR) study reporting that higher educational attainment drives mitigated DNAmGrimAA (McCartney et al. 2020), and with observational studies showing that DNAmGrimAA outperforms other epigenetic aging biomarkers in predicting mortality and age-related clinical phenotypes (McCrory et al. 2020a, Lu et al. 2019). We nevertheless acknowledge that there is limited MR evidence of a causal link between DNAmGrimAA and mortality (McCartney et al. 2020). However, these MR analyses may lack validity as they used weak genetic instruments and were not implemented for men and women separately.

In addition, we found that the potential path-specific contribution of epigenetic aging to the mortality excess rate, independent of unhealthy lifestyle and morbidities, was small. This result supported that the examined epigenetic aging biomarkers captured most of the biological embedding of unfavourable exposures (unhealthy lifestyles and/or morbidities) attendant on lower education, eventually leading to an increased mortality rate. As unhealthy lifestyle and morbidities are the target of policies promoted by the WHO to reduce the burden of premature mortality, our finding strenghtens the case for those policies and suggest they could additionally mitigate social inequalities in epigenetic aging, and eventually mortality (Laine et al. 2019). The small residual path-specific effect associated with epigenetic aging only, especially for biomarker DNAmGrimAA, might be attributed to unmeasured mediators such as those related to employment conditions or psychosocial resources, or to unmeasured unhealthy lifestyle factors such as poor quality diet. However, as lifestyle behaviours are known to cluster (Noble et al. 2015), our assessement of the joint mediation by four other unhealthy lifestyle factors potentially encompasses mediation by poor diet. Additionally, as DNAmGrimAA was calibrated on smoking pack years (Lu et al. 2019) among other phenotypes, the residual path-specific effect could be due to the inability of the examined smoking intermediate variable to capture mediation by smoking intensity. Finally, since the estimate for DNAmGrimAA had limited precision with compatibility intervals ranging from -4 to 19 excess deaths per 10,000 person-years and from 0.98 to 1.13 (hazard ratio), future studies with larger sample sizes and fine-grained lifestyle-related mediators are required to validate our results.

For women, the portion of educational inequalities in all-cause mortality explained by epigenetic aging biomarkers was small and negative on the absolute scale, and small and positive on the relative scale. These apparently contrasting results could be related to statistical constraints with disaggregating a small hazard difference into direct and indirect portions and the available sample size (Rudolph, Goin and Stuart 2020). On the other hand, as educational inequalities on the hazard ratio scale were not small, the little mediation on the relative scale may either suggest that educational inequalities in mortality in women get embedded mostly through alternative biological mechanisms – not involving changes in epigenetic aging based on DNA methylation in blood (McCrory et al. 2020b) –, or point to unaccounted potential heterogeneity by age at menopause, which has been shown to affect epigenetic aging biomarkers (Levine et al. 2016), or suggest that the epigenetic clock biomarkers should be calibrated separately for men and women to improve estimation of individual epigenetic age considering sex differences in aging (Yusipov et al. 2020).

Our work has limitations. Our findings may not be generalized to non-White populations as most participants were White and may suffer from differential mortality attrition as participants were 64 years old on average. The mediators – DNA methylation, lifestyle, and morbidities – were measured at a single time point, providing only a snapshot of the aging process. Measuring a mediator only once may bias its contribution (Oude Groeniger et al. 2017), so there is a need for future research to assess repeated measurements of DNA methylation in large samples of population-based cohorts.

Unmeasured confounding may have biased our estimates. However, we implemented several sensitivity analyses for potentially unmeasured confounding that supported our main findings. Additionally, we implemented a sensitivity analysis to accommodate potential measurement error affecting epigenetic aging biomarkers, and this analysis supported our main findings.

We modelled educational attainment with two levels, potentially limiting our ability to capture the entire education-related social stratification process, which may underestimate the effects on all-cause mortality. We postulated a hypothetical intervention on educational attainment to assess educational inequalities in mortality, however we did not operationalize how such an intervention could be formulated for policies. Aiming to raise educational attainment by increasing compulsory schooling or subsidizing secondary school may still have imperfect compliance and unpredictable effects on mortality. Therefore, our results cannot be interpreted as causal effects estimates of specific interventions aiming to reduce mortality by increasing educational attainment. Despite this, the effect estimates in the present study were robust and support DNA methylation-based biomarkers of epigenetic aging in leukocytes as surrogate endpoints to assess social inequalities in life expectancy. We acknowledge our results add one line of empirical evidence while do not provide definitive evidence, as with observational data we could not ascertain whether the differential changes in DNA methylation captured by the biomarkers had an active or passive role in driving mortality (Carmeli et al. 2021, Gutierrez-Arcelus et al. 2013). Future experimental and longitudinal studies in both animals and humans are needed to enable more robust causal claims about the role of DNA methylation in immune cells (Keenan and Allan 2019).

A major strength of this study was the use of a counterfactual mediation framework addressing issues that may ensue from traditional linear methods as the difference or product mediation methods (Baron and Kenny 1986). Specifically, IORW provides unbiased estimates when mediators are categorical, when there is an interaction between exposure and mediator(s), or when parameters are non-collapsible as for the Cox survival model (Daniel, Zhang and Farewell 2020). Additionally, IORW offers a sensitivity analysis to assess the impact of potentially unmeasured confounding of the mediator-outcome relationship (Nguyen et al. 2015). Lastly, IORW is parsimonious as it does not require to specify a model for the joint conditional density of mediators when estimating indirect effects by a bloc of mediators. Notably, we evaluated mediators *en-bloc* to assess the potential path-specific contribution of epigenetic aging independent of unhealthy lifestyle and morbidities. This joint mediation is advantageous as there is no need to specify a causal order among mediators (VanderWeele and Vansteelandt 2014, Laine et al. 2019), which would be challenging in our case as the directionality between and within unhealthy lifestyle factors and/or morbidities was questionable, and is robust to potentially unmeasured confounding of mediator-mediator relationships (VanderWeele and Vansteelandt 2014).

Additional strengths were the inclusion of multiple populations to make empirical evidence stronger, and the two-step approach for estimating effects in each cohort first, and then pooling these via a random effect meta-analysis. This was useful to consider potential inter-cohort differences determined by geography and period. Finally, we estimated effects of education on all-cause mortality on both absolute and relative scales, thereby providing a complete description of inequalities.

In conclusion, we have provided empirical evidence about the substantive role of blood DNA methylation-based epigenetic aging biomarkers in explaining educational inequalities in all-cause mortality for men. Whether epigenetic aging acts as a biomarker of longevity or as a causal molecular mechanism of healthy aging remains an open question. Importantly, our results further suggested that differential effects of unhealthy lifestyle and morbidities determine this epigenetic embedding of unfavorable educational exposures ultimately leading to increased mortality rates. It would be a step too far to suggest that designing policies only around such downstream pathways would be the most effective way to reduce social inequalities in premature mortality and population health, when in reality whole systems/whole population life-course approaches will be required (Lundberg 2020).

## Methods

### Cohort selection

Of the 18 cohorts included in LIFEPATH, an international consortium aiming to disentangle the biological pathways underlying social inequalities in health (Vineis et al. 2020b, Vineis et al. 2020a), only adult cohorts were retained for the present study. Of these, we further selected those with available DNA methylation and subsequent mortality follow-up. Based on these criteria, three cohorts were included, namely EPIC-IT (N = 1,545 in Italy), MCCS (N = 2,816 in Australia), and TILDA (N = 490 in Ireland). We also gathered data from five additional adult cohorts, namely ESTHER (N = 1,864 in Germany), KORA (N = 1,727 in Germany), MESA (N = 1,264 in United States of America), NICOLA (N = 1,981 in Northern Ireland, United Kingdom), and the Rotterdam Study (RS) (N = 1,420 in The Netherlands). These participants were all sub-samples of the corresponding cohorts. A full description of these cohorts and of the sampling design is presented as **Supplementary Material**.

### Exposure, mediators, and outcome

The proposed causal structures underlying our study are represented in **Figure 3**. We designed our causal modelling to evaluate path-specific mechanisms of educational inequalities in all-cause mortality (outcome). We focused on inequalities driven by educational attainment (exposure). The first causal model, depicted in **Figure 3A**, dissected the educational inequalities (total effect of the exposure) into a non-transmitted (direct) effect and a transmitted (indirect) effect by blood DNA methylation-based markers of epigenetic aging (mediator). In the second causal model, depicted in **Figure 3B**, the indirect effect via epigenetic aging biomarkers was disaggregated into a pathway through unhealthy lifestyle and morbidities (LM), and a pathway independent from the latter. This model assumed that epigenetic aging is downstream of LM mediators as supported by a recent Mendelian Randomization study (McCartney et al. 2020). By comparing the mediated effect via LM *en-bloc* and the joint mediated effect via LM *and* epigenetic aging *en-bloc*, we assessed whether epigenetic aging provides additional mediation compared to LM only, or whether the effect of education on all-cause mortality propagates through a pathway affecting epigenetic aging independently of unhealthy lifestyle and morbidities.

**Figure 3.**
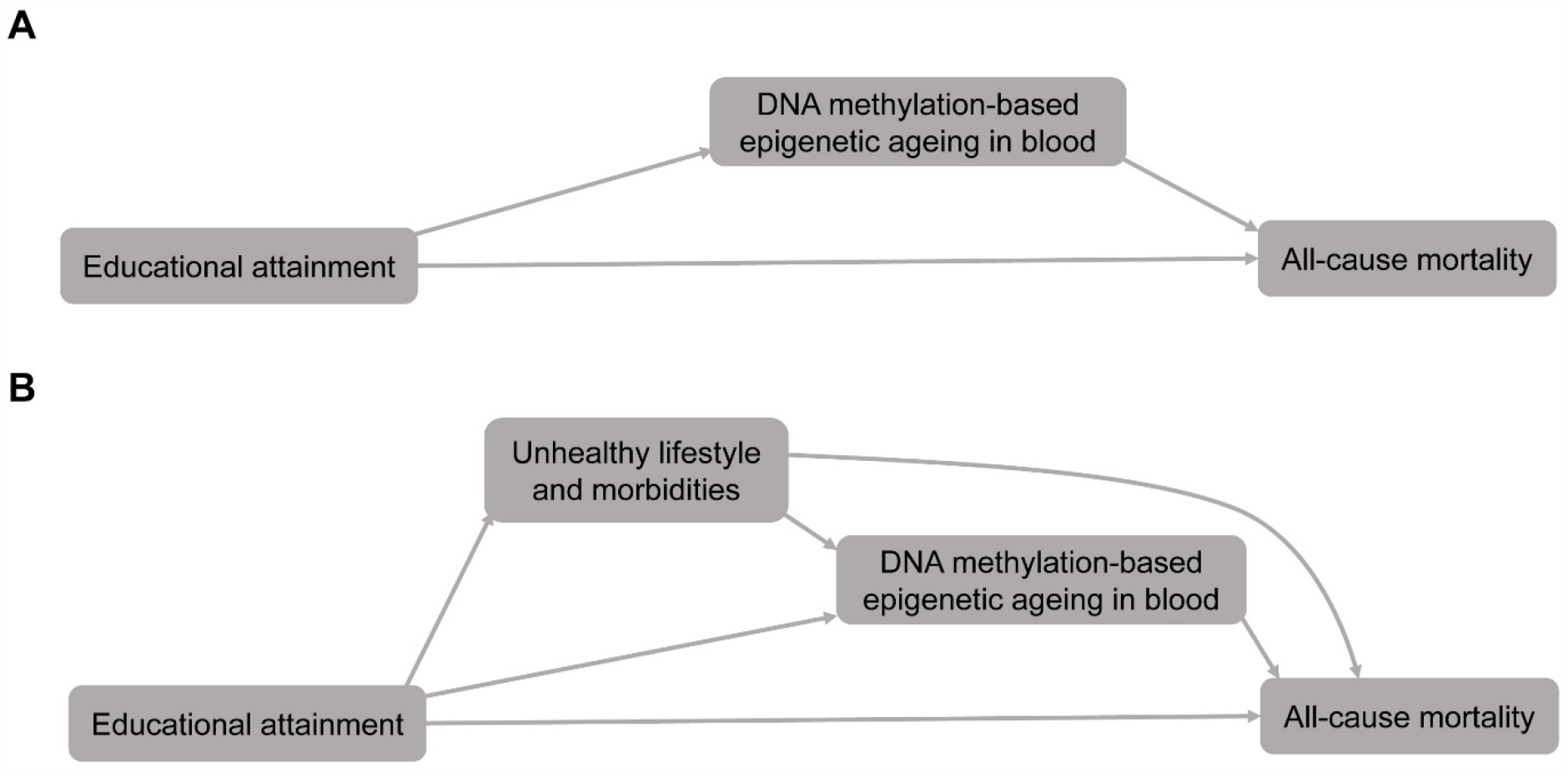
Causal structures. For the sake of simplicity, we do not include confounders (age and sex). Highest educational attainment represents the exposure and all-cause mortality the outcome. A) Adulthood DNA methylation-based epigenetic aging in blood is the investigated mediator. B) Adulthood unhealthy lifestyle (low physical activity, high alcohol intake, current smoking, and high body mass index), morbidities (diabetes and hypertension), and epigenetic aging in blood are the mediators. Unhealthy lifestyle and morbidities are considered mediators *en-bloc* and upstream of epigenetic aging.

Educational attainment was measured using the participants’ self-reported highest educational attainment. Answers were categorized as lower education for primary or lower secondary and higher for high-school or university degree. In **Supplementary Table 1** we report how educational attainment was harmonized across the included cohort studies.

Epigenetic aging in blood, unhealthy lifestyle and morbidities were included as mediators and measured at the same time, which we set as the baseline for mortality follow-up. In **Supplementary Table 2** we report the years of measurement of exposure and mediators for each cohort. Blood DNA methylation-based epigenetic age was assessed with four established epigenetic clock biomarkers: Horvath’s and Hannum’s DNAmAge (Horvath 2013, Hannum et al. 2013), calibrated on chronological age only, and Levine’s DNAmPhenoAge and Lu’s DNAmGrimAge (Horvath 2013, Hannum et al. 2013, Levine et al. 2018, Lu et al. 2019), calibrated on chronological age and factors associated with risk of mortality and incidence of aging-related diseases. We chose them as they have been commonly studied in the literature, were shown to be associated with both education and all-cause mortality in several populations, and although correlated may capture different aspects of epigenetic aging processes (Fiorito et al. 2019, Lu et al. 2019, McCartney et al. 2020, Wang et al. 2021). Elevated or mitigated epigenetic aging was measured as the residual (continuous and in years) from regressing these epigenetic clocks on chronological age, whereby positive/negative values of these residuals corresponded to elevated/mitigated epigenetic aging. Thereafter, we refer to these values as Horvath DNAmAA, Hannum DNAmAA, DNAmPhenoAA and DNAmGrimAA, respectively. Although these biomarkers are usually described as capturing accelerated or decelerated epigenetic aging, we find the words elevation and mitigation more appropriate for use in our study as they measure a difference rather than a rate of change. The proportion of leukocytes was estimated for each participant according to Houseman’s method (Houseman et al. 2012) and was used in sensitivity analyses. This method is recommended for cell-type heterogeneity adjustment of DNA methylation measured from blood (Zheng et al. 2017) and led to estimated proportions of T helper cells, cytotoxic T cells, natural killer cells, beta cells, monocytes, and granulocytes.

Unhealthy lifestyle comprised self-reported current smoking, high alcohol consumption, low level of leisure-time physical activity, and high body mass index (BMI) categorized into two or three-level variables (see **Supplementary Material**). Morbidities comprised hypertension (either self-reported or measured) and diabetes (either self-reported or measured). As highest education is generally attained in young adulthood whereas study participants were in their mid or late adulthood at baseline (see **Supplementary Table 2**), the potential for reverse causality was minimized. We chose these mediators as they are on the causal path from educational attainment to all-cause mortality (Ross and Wu 1995, Laine et al. 2019, Janke et al. 2020, Brunello et al. 2016), are targets of policies to reduce the burden of premature mortality (Stringhini et al. 2017), and are associated with the chosen epigenetic aging biomarkers (Fiorito et al. 2019, Lu et al. 2019).

Potential confounders were age at baseline (continuous), sex, and childhood socioeconomic conditions (available only in a subset of cohorts and included in a sensitivity analysis). In MESA, race/ethnicity was added as a potential confounder. Additionally, sex was modelled as an effect modifier, as previous observational studies have shown sex differences in educational inequalities in mortality (Galama et al. 2018, Laine et al. 2019).

The outcome was age-to-death from any cause, with age as the underlying timescale. This choice implicitly considered delayed entry or left truncation.

Detailed information about education, lifestyle, morbidities, DNA methylation, and mortality related variables in each cohort are reported in **Supplementary Material**.

### Statistical framework and analysis

We posited the counterfactual change of educational attainment from higher to lower to estimate the portion of educational inequalities in all-cause mortality explained by the chosen mediators. Within this framework, the educational inequalities in all-cause mortality correspond to the total effect of educational attainment, which can be decomposed into the direct effect – the portion of the inequalities not explained by a mediator –, and the indirect effect – the portion of the inequalities explained by a mediator (VanderWeele 2015). To estimate the effects of interest in our study – total and indirect – we used the inverse odds ratio weighting (IORW) method (Tchetgen Tchetgen 2013). Under certain identifying conditions (see **Supplementary Material**), IORW enables estimation of indirect and joint indirect effects (e.g. effect of exposure on outcome mediated by a bloc of mediators). The latter does not depend on a specific causal order among the mediators (VanderWeele and Vansteelandt 2014), which is useful in our study as specifying an order among unhealthy lifestyle and/or morbidities may be questionable. Additionally, contrary to linear path analysis, the IORW method allows estimation of a more general class of mediation models including those with mediators and/or outcome measured on a non-continuous scale (as in our study) and incorporates the potential interaction between exposure and mediator(s). The latter is useful as education and unhealthy lifestyle may interact in affecting mortality (Nordahl et al. 2014).

We assessed effects on the hazard difference and ratio scales, which correspond to an absolute and relative measure of inequalities, respectively. Both measures are useful in understanding and describing inequalities (Asada 2010). For the hazard difference, the total effect was defined as the difference in the number of deaths per 10,000 person-years had the exposure to education been changed from higher to lower (potentially counter to fact), while the indirect effect was the difference in the number of deaths per 10,000 person-years had the exposure to education been set to higher (potentially counter to fact) and the mediator(s) had changed from the value it would take at higher education to the value it would take at lower education (potentially counter to fact).

For analyses related to the model in **Figure 3A**, a chosen epigenetic aging biomarker was the only mediator, while for analyses related to the model in **Figure 3B**, seven mediators were considered. As we were interested in the portion of the total effect explained by epigenetic aging independent of unhealthy lifestyle and morbidities, we decomposed the joint indirect effect by the bloc of seven mediators into two path-specific indirect effects (Steen et al. 2017). Specifically, we decomposed the joint indirect effect by LM and epigenetic aging into the indirect effect by the bloc of mediators LM – capturing both pathways education→LM→mortality and education→LM→epigenetic aging→mortality –, and the indirect effect by epigenetic aging but not LM (see **Figure 3B**). We implemented a sequential mediation analysis to estimate the latter indirect effect as the difference between two joint indirect effects, the first being a joint indirect effect by all seven mediators and the second being a joint indirect effect by LM only.

All analyses were conducted on complete data. For the analyses based on the model in **Figure 3A**, we excluded 86 (0.7%) participants with missing information on education or vital status, while for the analyses based on the model in **Figure 3B**, we excluded 771 (5.9%) participants for missing information on education, vital status, unhealthy lifestyle, and morbidities.

For the main analyses, we applied a semiparametric additive hazard model (Martinussen and Sheike 2006, Nguyen et al. 2015) whereby hazards related to education and sex were held constant (see **Supplementary Material** for more details about the implementation of the IORW method). Effects on the hazard ratio scale were estimated by implementing a semiparametric multiplicative hazard model (Cox). In sensitivity analyses we evaluated potential deviations from the assumption of constant hazard differences and ratios (Stensrud and Hernán 2020).

For each cohort, we generated 5,000 bootstrap draws with replacement to derive standard errors for the total and indirect effects. Estimates from each cohort were pooled through a weighted inverse variance random effect meta-analytic model to consider potential heterogeneities across cohort studies (see **Supplementary Material**).

To assess the sensitivity of our findings to possible violations of the assumptions underlying the identification of the indirect effects, we estimated the indirect effect by epigenetic aging biomarkers (**Figure 3A**) i) when including childhood socioeconomic conditions as a potential confounder of the exposure-outcome, exposure-mediator, and mediator-outcome associations; ii) for various values of a bias function encoding unmeasured, potentially exposure-induced, confounding of the mediator-outcome association (Nguyen et al. 2015); and iii) when incorporating a non-differential measurement error of three years in epigenetic aging biomarkers. To assess the sensitivity of our findings to deviations from the assumption of constant hazard differences/ratios on age, we estimated the effects by incrementally right censoring survival age at 70, 75, 80, and 85 years. Finally, to assess the path-specific mediation by epigenetic aging biomarkers and not leukocyte composition in blood, we ran a sequential mediation analysis based on the model in **Supplementary Figure 6**. In **Supplementary Material** we provide detailed information about sensitivity analyses and childhood socioeconomic conditions.

## Data Availability

Cohort data are available upon request to the PI of each cohort study. Programming code that supports the findings of this study is available from the corresponding author upon request.

## References

Asada, Y. (2010) On the choice of absolute or relative inequality measures. Milbank Q, 88, 616–22; discussion 623-7.

Baron, R. M. & D. A. Kenny (1986) The moderator-mediator variable distinction in social psychological research: conceptual, strategic, and statistical considerations. J Pers Soc Psychol, 51, 1173–82.

Berger, E., R. Castagné, M. Chadeau-Hyam, M. Bochud, A. d’Errico, M. Gandini, M. Karimi, M. Kivimäki, V. Krogh, M. Marmot, S. Panico, M. Preisig, F. Ricceri, C. Sacerdote, A. Steptoe, S. Stringhini, R. Tumino, P. Vineis, C. Delpierre & M. Kelly-Irving (2019) Multi-cohort study identifies social determinants of systemic inflammation over the life course. Nat Commun, 10, 773.

Bosworth, B. (2018) Increasing Disparities in Mortality by Socioeconomic Status. Annu Rev Public Health, 39, 237–251.

Brunello, G., M. Fort, N. Schneeweis & R. Winter-Ebmer (2016) The Causal Effect of Education on Health: What is the Role of Health Behaviors? Health Econ, 25, 314–36.

Carmeli, C., Z. Kutalik, P. P. Mishra, E. Porcu, C. Delpierre, O. Delaneau, M. Kelly-Irving, M. Bochud, N. A. Dhayat, B. Ponte, M. Pruijm, G. Ehret, M. Kähönen, T. Lehtimäki, O. T. Raitakari, P. Vineis, M. Kivimäki, M. Chadeau-Hyam, E. Dermitzakis, N. Vuilleumier & S. Stringhini (2021) Gene regulation contributes to explain the impact of early life socioeconomic disadvantage on adult inflammatory levels in two cohort studies. Sci Rep, 11, 3100.

Castagné, R., C. Delpierre, M. Kelly-Irving, G. Campanella, F. Guida, V. Krogh, D. Palli, S. Panico, C. Sacerdote, R. Tumino, S. Kyrtopoulos, F. S. Hosnijeh, T. Lang, R. Vermeulen, P. Vineis, S. Stringhini & M. Chadeau-Hyam (2016) A life course approach to explore the biological embedding of socioeconomic position and social mobility through circulating inflammatory markers. Sci Rep, 6, 25170.

Castagné, R., V. Garès, M. Karimi, M. Chadeau-Hyam, P. Vineis, C. Delpierre, M. Kelly-Irving & L. Consortium (2018) Allostatic load and subsequent all-cause mortality: which biological markers drive the relationship? Findings from a UK birth cohort. Eur J Epidemiol, 33, 441–458.

Christiansen, L., A. Lenart, Q. Tan, J. W. Vaupel, A. Aviv, M. McGue & K. Christensen (2016) DNA methylation age is associated with mortality in a longitudinal Danish twin study. Aging Cell, 15, 149–54.

Daniel, R., J. Zhang & D. Farewell (2020) Making apples from oranges: comparing non-collapsible effect estimators and their standard errors after adjustment for different covariate sets. Biometrical Journal.

Dugué, P. A., J. K. Bassett, J. E. Joo, C. H. Jung, E. Ming Wong, M. Moreno-Betancur, D. Schmidt, E. Makalic, S. Li, G. Severi, A. M. Hodge, D. D. Buchanan, D. R. English, J. L. Hopper, M. C. Southey, G. G. Giles & R. L. Milne (2018) DNA methylation-based biological aging and cancer risk and survival: Pooled analysis of seven prospective studies. Int J Cancer, 142, 1611–1619.

Fiorito, G., C. McCrory, O. Robinson, C. Carmeli, C. O. Rosales, Y. Zhang, E. Colicino, P. A. Dugué, F. Artaud, G. J. McKay, A. Jeong, P. P. Mishra, T. H. Nøst, V. Krogh, S. Panico, C. Sacerdote, R. Tumino, D. Palli, G. Matullo, S. Guarrera, M. Gandini, M. Bochud, E. Dermitzakis, T. Muka, J. Schwartz, P. S. Vokonas, A. Just, A. M. Hodge, G. G. Giles, M. C. Southey, M. A. Hurme, I. Young, A. J. McKnight, S. Kunze, M. Waldenberger, A. Peters, L. Schwettmann, E. Lund, A. Baccarelli, R. L. Milne, R. A. Kenny, A. Elbaz, H. Brenner, F. Kee, T. Voortman, N. Probst-Hensch, T. Lehtimäki, P. Elliot, S. Stringhini, P. Vineis, S. Polidoro, B. Consortium & L. consortium (2019) Socioeconomic position, lifestyle habits and biomarkers of epigenetic aging: a multi-cohort analysis. Aging (Albany NY), 11, 2045–2070.

Galama, T., L.-M. A & H. van Kippersluis. 2018. The effect of education on health and mortality: A review of experimental and quasi-experimental evidence. Cambridge, MA: National Bureau of Economic Research: NBER Working Paper 24225.

Gallo, V., J. P. Mackenbach, M. Ezzati, G. Menvielle, A. E. Kunst, S. Rohrmann, R. Kaaks, B. Teucher, H. Boeing, M. M. Bergmann, A. Tjønneland, S. O. Dalton, K. Overvad, M. A. Daponte, L. Arriola, C. Navarro, A. B. Gurrea, K. T. Khaw, N. Wareham, T. Key, A. Naska, A. Trichopoulou, D. Trichopoulos, G. Masala, S. Panico, P. Contiero, R. Tumino, H. B. Bueno-de-Mesquita, P. D. Siersema, P. P. Peeters, S. Zackrisson, M. Almquist, S. Eriksson, G. Hallmans, G. Skeie, T. Braaten, E. Lund, A. K. Illner, T. Mouw, E. Riboli & P. Vineis (2012) Social inequalities and mortality in Europe--results from a large multi-national cohort. PLoS One, 7, e39013.

Gruenewald, T. L., A. S. Karlamangla, P. Hu, S. Stein-Merkin, C. Crandall, B. Koretz & T. E. Seeman (2012) History of socioeconomic disadvantage and allostatic load in later life. Soc Sci Med, 74, 75–83.

Gutierrez-Arcelus, M., T. Lappalainen, S. B. Montgomery, A. Buil, H. Ongen, A. Yurovsky, J. Bryois, T. Giger, L. Romano, A. Planchon, E. Falconnet, D. Bielser, M. Gagnebin, I. Padioleau, C. Borel, A. Letourneau, P. Makrythanasis, M. Guipponi, C. Gehrig, S. E. Antonarakis & E. T. Dermitzakis (2013) Passive and active DNA methylation and the interplay with genetic variation in gene regulation. Elife, 2, e00523.

Hamdi, N. R., S. C. South & R. F. Krueger (2016) Does education lower allostatic load? A co-twin control study. Brain Behav Immun, 56, 221–9.

Hannum, G., J. Guinney, L. Zhao, L. Zhang, G. Hughes, S. Sadda, B. Klotzle, M. Bibikova, J. B. Fan, Y. Gao, R. Deconde, M. Chen, I. Rajapakse, S. Friend, T. Ideker & K. Zhang (2013) Genome-wide methylation profiles reveal quantitative views of human aging rates. Mol Cell, 49, 359–367.

Hertzman, C. (2012) Putting the concept of biological embedding in historical perspective. Proc Natl Acad Sci U S A, 109 Suppl 2, 17160–7.

Hillary, R. F., A. J. Stevenson, D. L. McCartney, A. Campbell, R. M. Walker, D. M. Howard, C. W. Ritchie, S. Horvath, C. Hayward, A. M. McIntosh, D. J. Porteous, I. J. Deary, K. L. Evans & R. E. Marioni (2020) Epigenetic measures of ageing predict the prevalence and incidence of leading causes of death and disease burden. Clin Epigenetics, 12, 115.

Horvath, S. (2013) DNA methylation age of human tissues and cell types. Genome Biol, 14, R115.

Houseman, E. A., W. P. Accomando, D. C. Koestler, B. C. Christensen, C. J. Marsit, H. H. Nelson, J. K. Wiencke & K. T. Kelsey (2012) DNA methylation arrays as surrogate measures of cell mixture distribution. BMC Bioinformatics, 13, 86.

Janke, K., D. W. Johnston, C. Propper & M. A. Shields (2020) The causal effect of education on chronic health conditions in the UK. J Health Econ, 70, 102252.

Keenan, C. R. & R. S. Allan (2019) Epigenomic drivers of immune dysfunction in aging. Aging Cell, 18, e12878.

Laine, J. E., V. T. Baltar, S. Stringini, M. Gandini, M. Chadeau-Hyam, M. Kivimaki, G. Severi, V. Perduca, A. M. Hodge, P. A. Dugué, G. G. Giles, R. L. Milne, H. Barros, C. Sacerdote, V. Krogh, S. Panico, R. Tumino, M. Goldberg, M. Zins, C. Delpierre, P. Vineis & L. Consortium (2019) Reducing socio-economic inequalities in all-cause mortality: a counterfactual mediation approach. Int J Epidemiol.

Levine, M. E., A. T. Lu, B. H. Chen, D. G. Hernandez, A. B. Singleton, L. Ferrucci, S. Bandinelli, E. Salfati, J. E. Manson, A. Quach, C. D. Kusters, D. Kuh, A. Wong, A. E. Teschendorff, M. Widschwendter, B. R. Ritz, D. Absher, T. L. Assimes & S. Horvath (2016) Menopause accelerates biological aging. Proc Natl Acad Sci U S A, 113, 9327–32.

Levine, M. E., A. T. Lu, A. Quach, B. H. Chen, T. L. Assimes, S. Bandinelli, L. Hou, A. A. Baccarelli, J. D. Stewart, Y. Li, E. A. Whitsel, J. G. Wilson, A. P. Reiner, A. Aviv, K. Lohman, Y. Liu, L. Ferrucci & S. Horvath (2018) An epigenetic biomarker of aging for lifespan and healthspan. Aging (Albany NY), 10, 573–591.

Li, X., A. Ploner, Y. Wang, P. K. Magnusson, C. Reynolds, D. Finkel, N. L. Pedersen, J. Jylhävä & S. Hägg (2020) Longitudinal trajectories, correlations and mortality associations of nine biological ages across 20-years follow-up. Elife, 9.

Liu, Z., B. H. Chen, T. L. Assimes, L. Ferrucci, S. Horvath & M. E. Levine (2019) The role of epigenetic aging in education and racial/ethnic mortality disparities among older U.S. Women. Psychoneuroendocrinology, 104, 18–24.

Lu, A. T., A. Quach, J. G. Wilson, A. P. Reiner, A. Aviv, K. Raj, L. Hou, A. A. Baccarelli, Y. Li, J. D. Stewart, E. A. Whitsel, T. L. Assimes, L. Ferrucci & S. Horvath (2019) DNA methylation GrimAge strongly predicts lifespan and healthspan. Aging (Albany NY), 11, 303–327.

Lundberg, O. (2020) Is lack of causal evidence linking socioeconomic position with health an ‘inconvenient truth’? Eur J Public Health, 30, 619.

Mackenbach, J. P., I. Kulhánová, B. Artnik, M. Bopp, C. Borrell, T. Clemens, G. Costa, C. Dibben, R. Kalediene, O. Lundberg, P. Martikainen, G. Menvielle, O. Östergren, R. Prochorskas, M. Rodríguez-Sanz, B. H. Strand, C. W. Looman & R. de Gelder (2016) Changes in mortality inequalities over two decades: register based study of European countries. BMJ, 353, i1732.

Martinussen, T. & T. H. Sheike. 2006. Dynamic regression models for survival data. New York: Springer.

McCartney, D., J. Min, R. Richmond, A. Lu, M. Sobczyk, G. Davies & L. Broer (2020) Genome-wide association studies identify 137 loci for DNA methylation biomarkers of ageing. bioRxiV, https://doi.org/10.1101/2020.06.29.133702.

McCrory, C., G. Fiorito, B. Hernandez, S. Polidoro, A. M. O’Halloran, A. Hever, C. Ni Cheallaigh, A. T. Lu, S. Horvath, P. Vineis & R. A. Kenny (2020a) GrimAge outperforms other epigenetic clocks in the prediction of age-related clinical phenotypes and all-cause mortality. J Gerontol A Biol Sci Med Sci.

McCrory, C., G. Fiorito, S. McLoughlin, S. Polidoro, C. N. Cheallaigh, N. Bourke, P. Karisola, H. Alenius, P. Vineis, R. Layte & R. A. Kenny (2020b) Epigenetic Clocks and Allostatic Load Reveal Potential Sex-Specific Drivers of Biological Aging. J Gerontol A Biol Sci Med Sci, 75, 495–503.

Murtin, F., J. Mackenbach, D. Jasilionis & M. Mira d’Ercole. 2017. Inequalities in longevity by education in OECD countries: insights from new OECD estimates. Paris.

Nguyen, Q. C., T. L. Osypuk, N. M. Schmidt, M. M. Glymour & E. J. Tchetgen Tchetgen (2015) Practical guidance for conducting mediation analysis with multiple mediators using inverse odds ratio weighting. Am J Epidemiol, 181, 349–56.

Noble, N., C. Paul, H. Turon & C. Oldmeadow (2015) Which modifiable health risk behaviours are related? A systematic review of the clustering of Smoking, Nutrition, Alcohol and Physical activity (‘SNAP’) health risk factors. Prev Med, 81, 16–41.

Nordahl, H., T. Lange, M. Osler, F. Diderichsen, I. Andersen, E. Prescott, A. Tjønneland, B. L. Frederiksen & N. H. Rod (2014) Education and cause-specific mortality: the mediating role of differential exposure and vulnerability to behavioral risk factors. Epidemiology, 25, 389–96.

Oude Groeniger, J., C. B. Kamphuis, J. P. Mackenbach & F. J. van Lenthe (2017) Repeatedly measured material and behavioral factors changed the explanation of socioeconomic inequalities in all-cause mortality. J Clin Epidemiol, 91, 137–145.

Ross, C. E., R. K. Masters & R. A. Hummer (2012) Education and the gender gaps in health and mortality. Demography, 49, 1157–83.

Ross, C. E. & C.-l. Wu (1995) The links between education and health. 60, 719–745.

Rudolph, K. E., D. E. Goin & E. A. Stuart (2020) The Peril of Power: A Tutorial on Using Simulation to Better Understand When and How We Can Estimate Mediating Effects. Am J Epidemiol, 189, 1559–1567.

Schmitz, L. L., W. Zhao, S. M. Ratliff, J. Goodwin, J. Miao, Q. Lu, X. Guo, K. D. Taylor, J. Ding, Y. Liu, M. Levine & J. A. Smith. 2021. The Socioeconomic Gradient in Epigenetic Aging Clocks: Evidence from the Multi-Ethnic Study of Atherosclerosis and the Health and Retirement Study. medRxiv 2021.03.01.21252660.

Steen, J., T. Loeys, B. Moerkerke & S. Vansteelandt (2017) Flexible Mediation Analysis With Multiple Mediators. Am J Epidemiol, 186, 184–193.

Stensrud, M. J. & M. A. Hernán (2020) Why Test for Proportional Hazards? JAMA, 323, 1401–1402.

Stringhini, S., C. Carmeli, M. Jokela, M. Avendaño, P. Muennig, F. Guida, F. Ricceri, A. d’Errico, H. Barros, M. Bochud, M. Chadeau-Hyam, F. Clavel-Chapelon, G. Costa, C. Delpierre, S. Fraga, M. Goldberg, G. G. Giles, V. Krogh, M. Kelly-Irving, R. Layte, A. M. Lasserre, M. G. Marmot, M. Preisig, M. J. Shipley, P. Vollenweider, M. Zins, I. Kawachi, A. Steptoe, J. P. Mackenbach, P. Vineis, M. Kivimäki & L. consortium (2017) Socioeconomic status and the 25 × 25 risk factors as determinants of premature mortality: a multicohort study and meta-analysis of 1·7 million men and women. Lancet, 389, 1229–1237.

Tchetgen Tchetgen, E. J. (2013) Inverse odds ratio-weighted estimation for causal mediation analysis. Stat Med, 32, 4567–80.

van Hedel, K., F. J. van Lenthe, J. Oude Groeniger & J. P. Mackenbach (2018) What’s the difference? A gender perspective on understanding educational inequalities in all-cause and cause-specific mortality. BMC Public Health, 18, 1105.

VanderWeele, T. J. 2015. Explanation in Causal Inference: Methods for Mediation and Interaction. New York: Oxford University Press.

VanderWeele, T. J. & S. Vansteelandt (2014) Mediation Analysis with Multiple Mediators. Epidemiol Methods, 2, 95–115.

Vineis, P., M. Avendano-Pabon, H. Barros, M. Bartley, C. Carmeli, L. Carra, M. Chadeau-Hyam, G. Costa, C. Delpierre, A. D’Errico, S. Fraga, G. Giles, M. Goldberg, M. Kelly-Irving, M. Kivimaki, B. Lepage, T. Lang, R. Layte, F. MacGuire, J. P. Mackenbach, M. Marmot, C. McCrory, R. L. Milne, P. Muennig, W. Nusselder, D. Petrovic, S. Polidoro, F. Ricceri, O. Robinson, S. Stringhini & M. Zins (2020a) Special Report: The Biology of Inequalities in Health: The Lifepath Consortium. Front Public Health, 8, 118.

Vineis, P., C. Delpierre, R. Castagné, G. Fiorito, C. McCrory, M. Kivimaki, S. Stringhini, C. Carmeli & M. Kelly-Irving (2020b) Health inequalities: Embodied evidence across biological layers. Soc Sci Med, 246, 112781.

Wang, C., W. Ni, Y. Yao, A. Just, J. Heiss, Y. Wei, X. Gao, B. A. Coull, A. Kosheleva, A. A. Baccarelli, A. Peters & J. D. Schwartz (2021) DNA methylation-based biomarkers of age acceleration and all-cause death, myocardial infarction, stroke, and cancer in two cohorts: The NAS, and KORA F4. EBioMedicine, 63, 103151.

Yusipov, I., M. G. Bacalini, A. Kalyakulina, M. Krivonosov, C. Pirazzini, N. Gensous, F. Ravaioli, M. Milazzo, C. Giuliani, M. Vedunova, G. Fiorito, A. Gagliardi, S. Polidoro, P. Garagnani, M. Ivanchenko & C. Franceschi (2020) Age-related DNA methylation changes are sex-specific: a comprehensive assessment. Aging (Albany NY), 12, 24057–24080.

Zheng, S. C., S. Beck, A. E. Jaffe, D. C. Koestler, K. D. Hansen, A. E. Houseman, R. A. Irizarry & A. E. Teschendorff (2017) Correcting for cell-type heterogeneity in epigenome-wide association studies: revisiting previous analyses. Nat Methods, 14, 216–217.

