## Supplementary Material for "The mediating role of epigenetic clocks underlying educational inequalities in mortality: a multi-cohort study"

#### *Detailed cohort description*

EPIC-IT

The European Prospective Investigation into Cancer and Nutrition (EPIC) is a large European study on diet and cancer and has been previously described elsewhere (Riboli 2001, Palli et al. 2003). The Italian component of EPIC (EPIC-IT) recruited 47,749 adult volunteers (men and women) at five centres: Varese and Turin in northern Italy, Florence in central Italy and Naples and Ragusa in southern Italy. All participants signed an informed consent form and completed two questionnaires: one about dietary habit (food-frequency) and one about lifestyle, with information on education, socioeconomic status, occupation, history of previous illnesses and surgery, lifetime tobacco use and alcohol consumption and physical activity. EPIC-IT database records were linked to cancer and regional mortality registries after database quality control. All EPIC-IT centres except Naples are covered by population-based cancer registries. In Naples, follow-up information was collected from electronic hospital discharge records and also by periodic personal contact with participants. The study was approved by the ethical review boards of the International Agency for Research on Cancer, and of the collaborating institutions responsible for subject recruitment in each of the EPIC recruitment centres. All centres for EPIC Italy, except for Florence are represented in LIFEPATH.

*Education* level was collected at baseline via questionnaire as none, primary school, lower secondary school, vocational school, higher secondary school, BSc, and MSc.

*Paternal profession* during childhood was collected at baseline via questionnaire as farmer, retailer, not employed, clerical worker, professional and manager, unskilled worker, skilled worker. This was categorized into manual and non-manual profession to characterize paternal occupational position.

*DNA methylation measurements and pre-processing.* DNA from blood samples were extracted from buffy coats using the QIAsymphony DNA Midi Kit (Qiagen, Crawley, UK). Bisulphite conversion of 500 ng of each sample was performed using the EZ-96 DNA Methylation-Gold™ Kit according to the manufacturer's protocol (Zymo Research, Orange, CA). Then, bisulfite-converted DNA was used for hybridization on the Infinium HumanMethylation 450 BeadChip, following the Illumina Infinium HD Methylation protocol. Briefly, a whole genome amplification step was followed by enzymatic end-point fragmentation and hybridization to HumanMethylation 450 BeadChips at 48°C for 17 h, followed by single nucleotide extension. The incorporated nucleotides were labeled with biotin (ddCTP and ddGTP) and 2,4-dinitrophenol (DNP) (ddATP and ddTTP). After the extension step and staining, the BeadChip was washed and scanned using the Illumina HiScan SQ scanner. The intensities of the images were extracted using the GenomeStudio (v.2011.1) Methylation module (1.9.0) software, which normalizes within-sample data using different internal controls that are present on the HumanMethylation 450 BeadChip and internal background probes. The methylation score for each CpG was represented as a β-value according to the fluorescent intensity ratio representing any value between 0 (unmethylated) and 1 (completely methylated).

We extracted raw fluorescence intensities data from 'idat' files and performed background subtraction, colour bias correction, and fluorescence intensities normalization using in house developed R script described elsewhere (Campanella et al. 2015). Samples were excluded if the bisulphite conversion fluorescence intensity was low (i.e. less than 10,000 for both type-I and type-II probes). Methylation measures were set to missing if the detection p-value was higher than 0.01. Finally, samples and CpG sites with low call rate (<95%) were removed prior to the analysis, as long with probes with non-unimodal distribution (probably methQTL sites). Probe design bias was corrected using the beta-mixture quantile (BMIQ) normalization procedure implemented in the 'wateRmelon' R package (Pidsley et al. 2013). Known batch effect by plate and position on the Illumina Beadchip was removed before statistical analyses using the ComBat algorithm described by Johnson et al. (ComBat function in the "sva" R package) (Leek et al. 2012). Finally, the leukocyte composition in each sample was estimated according to Houseman's method (Houseman et al. 2012). Epigenetic clocks measures were derived according with the algorithms described in the Horvath online epigenetic age calculator (https://horvath.genetics.ucla.edu/html/dnamage/).

The sample with measured DNA methlyation included individuals from five nested case-control studies on breast, colon, and lung cancer, lymphomas, and myocardial infarction. Participants were sampled from the 47,749 participants of the EPIC Italy cohort and included 354 incident breast cancer cases, 169 incident colon cancer cases, 192 incident lung cancer cases, 72 incident lymphoma cases, 292 incident myocardial infarction cases and their 1,079 matched controls. Controls were individually matched on age (±5 years), sex, season of blood collection, center, and length of follow-up. We excluded cases whose diagnosis was made less than one year since blood draw (N=68). Overall, after DNA methylation data quality controls and sample filtering 1,803 EPIC Italian subjects were left. After excluding samples from Florence center, 1,545 participants were left to use in this study.

*Smoking* was self-reported and assessed at baseline via a questionnaire. Information was collected for specific time periods (at the age of 20, 30, 40 and 50 years, approximately) and number of pack years and cigarettes per day, including the cumulative length of periods of smoke cessation for both current and former smokers (except in Naples).

*Alcohol* consumption was self-reported and collected at baseline. Past alcohol consumption was assessed as the number of glasses and types of alcoholic beverage consumed per week at different ages (20, 30, 40, and 50 y of age) (except in Naples).

*Physical activity* was collected at baseline via questionnaire. Questions included information on physical activity related to individuals' current job (sedentary, mainly standing, manual work, very heavy manual work), floor climbing, walking, biking, gardening, and physical fitness. Information on these latter activities was collected relative to winter and summer months. Energy estimates were constructed for the amount of energy expenditure for each type of activity considering the age and weight of individual participants. A cumulative index was estimated to describe activity as sport at least once a week or not.

*Body mass index* was collected at baseline and defined by height and weight that were measured at enrolment with a standardized protocol. Body mass index was calculated as the ratio between weight in kg and squared height in metres.

*Diabetes* was assessed at baseline, at the enrolment interview on selected medical conditions.

*Hypertension* was assessed at baseline at the enrolment interview on selected medical conditions and defined as present if the participant was under medical treatment, or with a measured systolic blood pressure ≥140mmHg or diastolic blood pressure ≥90mmHg.

*Vital status* was ascertained from local demographic database via automated linkage from the population town offices and mortality registry. In LIFEPATH, follow-up of vital status is available up to 2014.

<u>ESTHER</u>

The ESTHER study is an ongoing population-based cohort study conducted in the federal state of Saarland, Germany (Raum et al. 2007). In brief, 9,949 older adults (50-75 years) were recruited by their general practitioners (GPs) during routine health check-ups (offered every two years to people older than 35 years in the German healthcare system) between 2000 and

2002 and followed up thereafter. During the baseline enrolment, epidemiological data (including socio-demographic characteristics, lifestyle factors, and history of major diseases) were collected via a standardized self-administered questionnaire completed by participants and via additional reports from participants' GPs, and biological samples (blood, stool, urine) were obtained and stored at −80 °C. The study was approved by the ethics committees of the University of Heidelberg and of the Medical Association of Saarland. All participants provided written informed consent.

*Education* level was collected at baseline via questionnaire as no education, Hauptschule, Realschule, Mittlere reife, Fachhochschulreife, Abitur.

*DNA methylation measurements and pre-processing*. DNAm in whole blood was quantified using the Infinium HumanMethylation450K BeadChip (Illumina, Inc, San Diego, CA, USA) Details of methylation analysis in the ESTHER study have been reported previously (Zhang et al. 2016). According to the manufacturer's protocol, data were normalized to internal controls provided by Illumina (Illumina normalization). In data pre-processing, probes with detection *P*-value>0.01, with missing values>10%, probes targeting the sex chromosomes, cross-reactive probes and polymorphic CpGs55 were excluded, leaving 430,363 CpGs for genome-wide screening. In the KORA study, data were pre-processed following the pipeline of Lehne (Lehne et al. 2015), probes with detection *P*-value (1−*P*-value computed from the background model characterizing the probability that the target sequence signal was distinguishable from the negative controls) >0.01 and missing values >5% were removed, and quantile normalization was applied following stratification of the probe categories into six types, based on probe type and colour channel, using the R package limma (Smyth 2005). Leukocyte composition was estimated using the algorithms of Houseman *et al*. (Houseman et al. 2012). Epigenetic clocks were derived using the Horvath online epigenetic age calculator (https://horvath.genetics.ucla.edu/html/dnamage).

Two subsets of ESTHER participants were selected for DNA methylation assessment in the baseline blood samples: Subset I consisted of 1,000 participants consecutively enrolled during the first 3 months of recruitment; Subset II consisted of 864 participants selected for a case-control design for mortality analysis (Zhang et al. 2017). In mediation analyses, we pooled the two subsets at individual level and included an indicator variable as covariate to consider the potential differences between the two sub-cohorts.

*Smoking* was self-reported and assessed at baseline via a questionnaire and coded as 'Never', 'Former', or 'Current' smoker.

*Alcohol* consumption was self-reported and collected at baseline. Grams of daily alcohol consumption were coded as abstainer, moderate, and high alcohol intake following WHO standards.

*Physical activity* was collected at baseline via questionnaire. Participants who responded to practice ≥2h of vigorous or light physical activity per week were classified as active, and not active otherwise.

*Body mass index* was collected at baseline and defined by height and weight that were measured at enrolment with a standardized protocol. Body mass index was calculated as the ratio between weight in kg and squared height in metres.

*Diabetes* was assessed at baseline and determined from self-reported use of glucose lowering drugs, or self-reported physician diagnosis, or measured HbA1c levels ≥6.5%, or fasting glucose levels ≥126 mg/dL, or non-fasting glucose levels ≥200 mg/dL.

*Hypertension* was assessed at baseline and determined from self-reported use of anti-hypertensive treatment, or self-reported physician diagnosis, or measured systolic blood pressure ≥140mmHg or diastolic blood pressure ≥90mmHg.

*Vital status* was ascertained from local demographic database via automated linkage from the population town offices and mortality registry. Follow-up of vital status was available up to 2016.

MCCS

The Melbourne Collaborative Cohort Study (MCCS) has been fully described in a cohort profile(Milne et al. 2017). It is a prospective cohort study of 41,513 participants living in Melbourne, Australia. Caucasian volunteers aged between 40 and 69 years were recruited mainly from the Victorian Electoral Enrolment Register (enrolment to vote is compulsory in Australia) and the Melbourne metropolitan telephone directory. At baseline (1990–1994), participants attended clinics where demographic, lifestyle and dietary information were collected, and anthropometric measurements were performed. The Cancer Council Victoria's Human Research Ethics Committee approved the study protocol.

*Education* was derived at baseline. Education was collected as an ordinal variable, as never attended school, attended some primary school, completed primary school, attended some high/technical school, obtained other qualification (e.g. trade certificate), completed high school or technical school, completed a tertiary degree or diploma, completed some study towards a tertiary degree or diploma.

*DNA* samples used for the present analysis were extracted from peripheral blood drawn at the time of recruitment (1990–1994). For the majority (70%) of participants, the DNA source was dried blood spots collected onto Guthrie Card Diagnostic Cellulose filter paper (Whatman plc, Kent, United Kingdom) and stored in airtight containers at room temperature. The other sources of DNA were peripheral blood mononuclear cells and buffy coats stored at −80°C for 28% and 2% participants, respectively. The study sample comprised Melbourne Collaborative Cohort

Study participants selected as controls in nested case-control studies of breast, colorectal, kidney, lung, prostate, or urothelial cancer or mature B-cell malignancies (Severi et al. 2014, Dugué et al. 2016, Wong Doo et al. 2016, Baglietto et al. 2017). Controls had been individually matched to cases on age (they had to be free of cancer at an age within 1 year of the age at diagnosis of the corresponding case), sex, country of birth, and blood DNA source (dried blood spot, peripheral blood mononuclear cells, or buffy coat). For all but the colorectal cancer study, controls were matched to cases on year of birth. For the lung cancer study, controls were additionally matched on smoking status at the time of blood collection. Methods relating to DNA extraction and bisulphite conversion and DNA methylation data processing have been described in detail elsewhere (Dugué et al. 2018). Briefly, a total of 200 ng of bisulphite-converted DNA was whole-genome amplified and hybridised onto the BeadChips. Bisulphite conversion and quantification of DNA methylation with the Infinium HumanMethylation450 BeadChip were performed according to manufacturer's instructions. DNA methylation data were pre-processed and normalized using in house developed R script described elsewhere (Campanella et al. 2015). Samples were excluded (N = 124) if the bisulphite conversion control fluorescence intensity was less than 10,000 for both type I and type II probes. DNA methylation levels were expressed as the ratio of the intensities of methylated cytosines over the total intensities (β values). Each methylation measure was set to missing if the detection p-value was greater than 0.01; probes and samples were excluded if the total call rate was less than 95% (N = 255). Additionally, the set of cross-reactive and/or polymorphic probes with minor allele frequency higher than 0.01 in Europeans was excluded due to the low reliability of methylation measures. Forty-six samples were excluded because the estimated sex did not match the sex reported by the participant. Finally, a total of N = 2,816 participants had DNA methylation data available.  Known batch effect by plate and position on the Illumina Beadchip was removed before statistical analyses and epigenetic clock computation using the ComBat algorithm described by Johnson et al. (ComBat function in the "sva" R package) (Leek et al. 2012). Finally, the leukocyte composition in each sample was estimated according to Houseman's method (Houseman et al. 2012).

*Smoking* was self-reported and assessed at baseline via a questionnaire and coded as 'Never', 'Former', or 'Current' smoker.

*Alcohol* was self-reported and assessed at baseline via a questionnaire that asked about beverage specific (beer, wine, spirits) intake by unit and frequency for the current decade of life.

*Physical activity* was measured form questions relating to frequency of walking, vigorous exercise (defined as exercise that induced sweat or feelings of out of breath, including swimming, tennis, netball, athletics, and running), and less vigorous exercise (defined as exercise that did induce sweat or feelings of out of breath, including bike riding, dancing, etc) over the last 6 months were asked. The reported frequency for each question was coded as follows: 0 (none), 1.5 (one or two times per week), and 4 (three or more times per week). Walking and less vigorous exercise frequencies were added together along with two times the frequency of vigorous exercise to generate a physical activity score for each person, in minutes per week. Participants with less than 75 min/week were classified as having low physical activity, and those with more than 75 min/week as having high physical activity.

*Body mass index* was assessed at baseline and follow-up and based off of height and weight were measured at enrolment with a standardized protocol and body mass index (BMI calculated as the ratio between weight in kg and squared height in metres).

*Diabetes* was assessed at baseline and follow-up and based off of self-reported diabetes status (yes/no) and status based on blood glucose measured at baseline and self-report. For incident cases of type 2 diabetes, 76% had their diagnosis confirmed by their doctor.

*Hypertension* was assessed at baseline and follow-up via self-reported high blood pressure, self-reported anti-hypertensive medication, and (average from two measures) value of systolic blood pressure≥140 mmHg or diastolic blood pressure≥90 mmHg.

*Vitality* was identified by regular matching of the Melbourne Collaborative Cohort Study to cancer registries and death indices. Non-cancer, non-fatal health events were captured by following up the cohort with a mailed questionnaire and by telephone. Follow-up of vital status was available up to 2015. Participants were considered to be at risk from their index date (the date on which they reached the age at which their matched case in the corresponding nested case-control study was diagnosed with cancer) onward in order to avoid immortal time bias, as they had to be alive at this date.

KORA

The KORA project (Cooperative Health Research in the Region of Augsburg) consists of four independent cross-sectional surveys (S1–S4) starting between 1984 and 2001, and their respective follow-up studies (Holle et al. 2005). All participants were from the city or region of Augsburg. All participants were living in Germany and all were of European origin. For the present study, only participants in the KORA-F4 study (2006-2008) for which a DNA methylation measurement was available were included.

Genome-wide DNA methylation measurement at 485,577 genomic sites from whole-blood was performed using the Infinium HumanMethylation450K BeadChip® (Illumina, Inc., CA, USA) in 1,802 KORA F4 samples. These participants were chosen randomly stratified for age groups, where older participants were oversampled compared to younger ones. Sample preparation and measurement have been described previously (Zeilinger et al. 2013, Wilson et al. 2017). Methylated and unmethylated signal intensities were obtained for each CpG site, which were then converted to β-values, the ratio of the methylated signal intensity divided by the overall signal intensity. DNA methylation data were pre-processed following the CPACOR pipeline (Lehne et al. 2015). First, 65 probes that represent SNPs were excluded. Second, background correction was performed using the R package minfi, version 1.6.0 (Aryee et al. 2014). Third, detection p-values were defined as the probability of a signal being detected above the background signal level, as estimated from negative control probes. Consequently, signals with detection p-values $\geq 0.01$ were removed, as they indicate putatively unreliable signals. Similarly, signals summarized from less than three functional beads on the chip were characterized as potentially unreliable and removed from the data set. Observations with less than 95% of CpG sites providing reliable signals (75 in KORA F4) were excluded, resulting in 1,727 samples overall.

Data were normalized using quantile normalization on the raw signal intensities. Beta-mixture quantile normalization was performed on a stratification of the probe categories into 6 types, based on probe type and colour channel, using the R package limma, version 3.16.5 (Smyth 2005). These data were then uploaded to Horvath's online calculator to generate the various clock estimates.

*Smoking* was self-reported and assessed at baseline via a questionnaire. Regular and irregular smoker were coded as 'current' smoker, the remaining individuals were divided in 'former' smoker and 'never' smoker.

*Alcohol* consumption was collected at baseline based on average self-reported consumption (in g/day). Participants were asked how much beer, wine, and spirits they consumed on the previous weekday and weekend and were divided in 'abstainer', 'moderate' and 'high' consumption.

*Physical activity* was collected at baseline via questionnaire. Participants were asked how often they practice sports in summer/winter, with possible answers (1) regularly more than 2 hours a week, (2) regularly 1 to 2 hours a week, (3) less than 1 hours per week and (4) no sport in summer/winter. If the combined sum score was lower than 5, then participants were classified as 'active', and 'not active' otherwise.

*Body mass index* was collected at baseline and defined by height and weight, which were measured at enrolment with a standardized protocol. Body mass index was calculated as the ratio between weight in kg and squared height in metres.

*Diabetes* was assessed at baseline. The definition is based on a self-reported diabetes diagnosis, which was validated by the participant´s general practitioner, or antidiabetic medication intake.

*Hypertension* was assessed at baseline and defined as present if the participant was under medical treatment for hypertension, or with a measured systolic blood pressure ≥140mmHg or diastolic blood pressure ≥90mmHg.

*Vital status* was ascertained from local demographic database via automated linkage from the population town offices and mortality registry. Follow-up of vital status was available up to 2016.

MESA

The *Multi-Ethnic Study of Atherosclerosis* (MESA) enrolled 6,814 participants aged 45–84 free of cardiovascular diseases at baseline from July 2000 to July 2002 at six field centres (Baltimore, MD; Chicago, Illinois; Los Angeles, CA; New York, NY; St. Paul, MN, and Winston-Salem, NC) with ongoing follow-up. The MESA cohort is a diverse, population-based sample of 38% white, 28% black, 22% Hispanic, and 12% Asian participants, of whom half are women. Study participants underwent extensive physical exams to determine subclinical cardiovascular diseases and questionnaires to obtain information on sociodemographic, lifestyle, and psychosocial factors. The MESA study protocol was approved by the Institutional Review Boards (IRBs) at Johns Hopkins Medical Institutions, University of Minnesota, Columbia University Medical Center, Wake Forest University Health Sciences, and University of Washington. Analysis for this study was approved by the University of Michigan IRB. All participants signed informed consent.

*Education* was assessed at baseline via questionnaire as no schooling; grades 1-8, grades 9-11, completed high school/GED, some college but no degree, technical school certificate, associate degree, bachelor's degree, and graduate or professional school.

*Race/ethnicity* was self-reported at baseline as White, Asian, Black and Hispanic.

*Maternal and paternal education* were collected at the second examination between 2002 and 2004. Participants reported whether their mother/father had no schooling, some schooling but did not complete high school, high school degree, some college, college degree, graduate or professional school.

*DNA methylation* was measured within the MESA Epigenomics and Transcriptomics Study. Genome-wide methylomic and transcriptomic profiles of CD14+ purified monocytes were obtained from 1,264 randomly selected MESA participants from four MESA field centers (Baltimore, MD; New York, NY; St. Paul, MN; and Winston-Salem, NC) at the 5$^{th}$ examination (April 2010-February 2012) (Liu et al. 2013). These participants were 47% white. A detailed description of the data extraction and processing procedures used for DNA methylation can be found in Liu(Liu et al. 2013). Briefly, blood was drawn in the morning after a 12 hour fast. Monocytes were isolated using AutoMACs automated magnetic separation units (Miltenyi Biotec, Bergisch Gladbach, Germany) and were consistently >90% pure. Samples were plated using a stratified random sampling technique to reduce bias due to batch, chip, and position effects. Methylation was measured using the Illumina HumanMethylation450 BeadChip, and bead-level data were summarized in GenomeStudio. Quantile normalization was performed using the *lumi* package (Du, Kibbe and Lin 2008). Quality control measures included checks for sex and race/ethnicity mismatches and outlier identification by multidimensional scaling plots. Criteria for elimination included: 'detected' methylation levels in <90% of MESA samples (using a detection p-value threshold of 0.05) or overlap with a repetitive region. 65 probes that assay highly polymorphic single nucleotide polymorphisms (SNPs) rather than methylation were also excluded (Pidsley et al. 2013). DNA methylation clocks were calculated using an R script, with the exception of DNAmGrimAge, which was calculated using the Horvath online DNA Methylation Age Calculator (http://dnamage.genetics.ucla.edu/new).

*Smoking* was self-reported and assessed at the 5$^{th}$ examination via a questionnaire.

*Alcohol* consumption was self-reported and collected at the 5$^{th}$ examination. Reported weekly consumption of red/white glasses, beer and liquor or mixed drinks were converted into alcohol units and then categorized according to WHO standards.

*Physical activity* was collected at the 5$^{th}$ examination via questionnaire- Self-reported total intentional exercise encompassing walking for exercise, dance, sports, individual activities, and conditioning were converted in MET (min/week). Participants scoring ≥600 MET were categorized as physically active, and not physically active otherwise.

*Body mass index* was collected at the 5$^{th}$ examination and defined by height and weight that were measured at enrolment with a standardized protocol. Body mass index was calculated as the ratio between weight in kg and squared height in metres.

*Diabetes* was assessed at the 5th examination, and established following the 2003 diagnostic criteria of American Diabetes Association about impaired fasting glucose, that encompassed self-reported anti-diabetic treatment and fasting glucose measurement.

*Hypertension* was assessed at the 5th examination defined as present if averaged second and third readings of systolic blood pressure was ≥140mmHg or of diastolic blood pressure was ≥90mmHg.

*Vital status* was ascertained from local demographic database via automated linkage from the population town offices and mortality registry. Follow-up of vital status was available up to 2019.

NICOLA

The *Northern Ireland Cohort for the Longitudinal Study of Ageing* (NICOLA) is a longitudinal cohort representative of the non-institutionalized population of Northern Ireland 50 years and older (n=8,500) (Burns et al. 2017). The study which was established in 2013 has three main components: a computer aided personal interview (CAPI), a self-completion questionnaire and health assessment. Dietary intake was also assessed by a food frequency questionnaire. The CAPI was extensive in scope and included assessment of demographic, social and health-related factors. Measures of cardiovascular, physical, cognitive and visual function were determined, and a biobank of biological samples collected.

*Education* was self-reported at baseline via questionnaire as none, some primary (not complete), primary or equivalent, GCSE/Intermediate/junior/group certificate or equivalent, A-level/Leaving certificate or equivalent, diploma/certificate, primary degree, postgraduate/higher degree.

*Financial situation before age 14* was self-reported at baseline as poor, about average, pretty well off financially.

*DNA methylation measurement*

All the study participants that attended the health assessment and provided a venous blood sample were analysed for DNA methylation profiles. DNAm measurement was performed from blood cells using the Illumina850K BeadChip array, using standard protocols suggested by the manufacturers. Raw methylation data were pre-processed and analysed as previously described for the EPIC Italy cohort. Epigenetic clocks measures were derived as previously described for the EPIC Italy cohort.

*Smoking* was self-reported and assessed at baseline via a questionnaire.

*Alcohol* consumption was self-reported and collected at baseline as total number of alcohol units per week.

*Physical activity* was collected at baseline via questionnaire. The sum of vigorous and moderate leisure physical activities and walking activities were estimated as MET (min/week). Participants scoring ≥600 MET were categorized as physically active, and not physically active otherwise.

*Body mass index* was collected at baseline and defined by height and weight that were measured at enrolment with a standardized protocol. Body mass index was calculated as the ratio between weight in kg and squared height in metres.

*Diabetes* was assessed at baseline as self-reported intake of anti-diabetic drugs or measured HbA1c levels ≥6.5%.

*Hypertension* was assessed at baseline as self-reported intake of anti-hypertensive drugs, physician diagnosis, or measured systolic blood pressure ≥140mmHg or diastolic blood pressure ≥90mmHg (average of last two readings).

*Vital status* was ascertained from local demographic database via automated linkage from the Business Services Organisation (BSO), an agency of the Department of Health, which uses information from the Northern Ireland Registrar General together with data from registrations and pensions to identify all deaths in Northern Ireland. Follow-up of vital status was available up to 31st May 2020.

RS

The *Rotterdam Study* (RS) is a prospective population-based study started in 1989. It is composed of residents of the neighbourhood of Ommoord, Rotterdam, the Netherlands, aged 45 years and over. Each participant gave an informed consent and the study was approved by the medical ethics committee of the Erasmus University Medical Center, Rotterdam, the Netherlands.

Data on socio-economic status, diet and lifestyle factors were assessed by standardized questionnaires, measured anthropometric data and blood samples collected for DNA extraction.

*Education* was categorized into three groups: i) ‘low’ = primary school or lower; ii) ‘medium’ = secondary school; iii) ‘high’ = university degree or higher.

DNA for methylation measurement was extracted from whole blood (stored in EDTA tubes) by standardized salting out methods, from a randomly selected sample of 1,471 individuals of RS second (RSII) and third (RSIII) sub-cohorts. Purified DNA was bisulfite-converted using the Zymo EZ DNA Methylation kit (Zymo Research Corporation) Genome-wide DNA methylation was measured in genomic DNA using the Illumina Infinium HumanMethylation 450 BeadChip (HM450) (Illumina, CA, USA), according to manufacturer's protocols. Preparation and normalization of this array data was performed according to the CPACOR workflow (Lehne et al. 2015). The idat files were read using the minfi package. Samples showing inadequate hybridization, incomplete bisulfite treatment and gender swaps were excluded. Samples were excluded if signal detection rate across probes was < 95%, leaving 1,420 individuals. CpG methylation proportion was reported as a normalized β-value ranging from 0 to 1, where 1 represents 100% methylation. β values being outside 1.5 times the interquartile range per probe were set to missing. Epigenetic clocks were estimated as described for EPIC-IT.

*Smoking* was categorized as 'never', 'former' and 'current' smokers based on self-reported information.

*Physical activity level* was measured with a self-administrated LASA Physical Activity questionnaire (LAPAQ). Later, the intensity of the reported activities was quantified using the metabolic equivalent of task (MET) hours per week, and then categorized in three groups: i) sedentary: <10 MET hours/week; ii) moderately active: 10–40 MET hours/week and iii) active: > 40 MET hours/week.

*Alcohol* consumption was collected using a validated semiquantitative food-frequency questionnaire. Frequency and amount of alcohol consumed was reported.

*Body mass index* (BMI) was calculated as the ratio between weight in kg and squared height in meters, and then categorized in three groups: i) normal weight: BMI ≤ 25; ii) overweight: 25 < BMI ≤ 30; iii) obese = BMI ≥ 30

*Type 2 diabetes* cases were defined as the presence of either fasting serum glucose level ≥7.0 mmol/L, physician diagnose or the use of glucose-lowering medication according to pharmacy records.

*Hypertension* was defined as an average value (of two measurements) of ≥140 mmHg for systolic- and ≥90 mmHg for diastolic blood pressure, or the use of antihypertensive medication, according to pharmacy records.

*Vital status* was obtained from the municipal health authorities in Rotterdam. Follow-up of vital status was available up to May 2018.

TILDA

*The Irish Longitudinal Study on Ageing (TILDA)* is a large prospective cohort study examining the social, economic and health circumstances of 8,175 community-dwelling older adults aged 50 years and over resident in the Republic of Ireland. The sample was generated using a 3-stage selection process and the Irish Geodirectory as the sampling frame. The Irish Geodirectory is a comprehensive listing of all addresses in the Republic of Ireland, which is compiled by the national post service and ordnance survey Ireland. Subdivisions of district electoral divisions pre-stratified by socio-economic status, age, and geographical location, served as the primary sampling units. The second stage involved the selection of a random sample of 40 addresses from within each PSU resulting in an initial sample of 25,600 addresses. The third stage involved the recruitment of all members of the household aged 50 years and over. Consequently, the response rate was defined as the proportion of households including an eligible participant from whom an interview was successfully obtained. A response rate of 62% was achieved at the household level. There were three components to the survey. Respondents completed a computer-assisted personal interview and a separate self-completion paper and pencil module which collected information that was considered sensitive. All participants were invited to undergo an independent health assessment at one of two national centers using trained nursing staff. Blood samples were taken during the clinical assessment with the consent of participants. A more detailed exposition of study design, sample selection and protocol are available elsewhere (Whelan and Savva 2013).

*Education* was self-reported at baseline via questionnaire as none, some primary (not complete), primary or equivalent, intermediate/junior/group certificate or equivalent, leaving certificate or equivalent, diploma/certificate, primary degree, postgraduate/higher degree.

*Paternal profession* during childhood was collected at baseline via questionnaire as farmer, unskilled, semi-skilled, non-manual, managerial and technical, professional worker. This was categorized into manual and non-manual profession to characterize paternal occupational position.

*DNA methylation and pre-processing*. The study sample included 500 healthy individuals: 125 for each of the four SES classes: stable professional, any downward mobility, any upward mobility, and stable unskilled. Buffy coat or peripheral blood mononuclear cells samples were available for all the individuals and extracted at baseline. Overall, after DNA methylation data quality controls and sample filtering, 490 subjects were retained. DNA samples were extracted from buffy coats using the QIAGEN GENTRA AUTOPURE LS (Qiagen, Crawley, UK). Bisulphite conversion of 500 ng of each sample was performed using the EZ DNA Methylation-Lightning™ Kit according to the manufacturer's protocol (Zymo Research, Orange, CA). Then, bisulfite-converted DNA was used for hybridization on the Infinium HumanMethylation 850k BeadChip, following the Illumina Infinium HD Methylation protocol. Briefly, a whole genome amplification step was followed by enzymatic end-point fragmentation and hybridization to HumanMethylation EPIC Chip at 48°C for 17 h, followed by single nucleotide extension. The incorporated nucleotides were labeled with biotin (ddCTP and ddGTP) and 2,4-dinitrophenol

(DNP) (ddATP and ddTTP). After the extension step and staining, the BeadChip was washed and scanned using the Illumina HiScan SQ scanner. The intensities of the images were extracted using the GenomeStudio (v.2011.1) Methylation module (1.9.0) software, which normalizes within-sample data using different internal controls that are present on the HumanMethylation 850k BeadChip and internal background probes. The methylation score for each CpG was represented as a β-value according to the fluorescent intensity ratio representing any value between 0 (unmethylated) and 1 (completely methylated). DNA methylation data were pre-processed and normalized using in-house software written for the R statistical computing environment, including background and color bias correction, quantile normalization, and BMIQ procedure to remove type I/type II probes bias, as described elsewhere (Teschendorff et al. 2013). DNAm levels were expressed as the ratio of the intensities of methylated cytosines over the total intensities (β values). Samples were excluded (N=4) if the bisulfite conversion control fluorescence intensity was less than 10,000 for both type I and type II probes. Methylation measures were set to missing if the detection p-value was greater than 0.01. No samples were excluded because of total call rate lower than 95%. Finally, samples (N=6) were excluded if the predicted sex (based on chromosome X methylation) did not match that self-reported. Epigenetic clocks measures were derived as previously described for EPIC-IT.

*Smoking* was self-reported and assessed at baseline via a questionnaire.

*Alcohol* consumption was self-reported and collected at baseline. A derived variable describing average number of drinks per week was obtained.

*Physical activity* was collected at baseline via the International Physical Activity Questionnaire (IPAQ) short form scoring protocol. Participants scoring moderate or high exercise intensity were categorized as physically active, while participants scoring low exercise intensity were assigned to being not physically active.

*Body mass index* was collected at baseline and defined by height and weight that were measured at enrolment with a standardized protocol. Body mass index was calculated as the ratio between weight in kg and squared height in metres.

*Diabetes* was assessed at baseline via self-reported physician diagnosis or HbA1c values ≥6.5%.

*Hypertension* was assessed at baseline via self-reported anti-hypertensive medications or a measured systolic blood pressure ≥140mmHg or diastolic blood pressure ≥90mmHg.

*Vital status* was available from two sources: (i) through data linkage to the General Registrar's Office (GRO) National Death Registry which was done in March 2017; and (ii) via an End of Life (EOL) interview for deaths occurring subsequent to administrative record linkage, which was conducted with the respondent's next of kin (if available). Follow-up of vital status was available up to December 2018. In 11 individuals for whom date of death was not available, their date of death was estimated as last interview observation + half of the inter-wave interval (approximately 1 year).

***Childhood socioeconomic conditions***

Proxies of socioeconomic conditions in childhood were available in four cohort studies and were included as a potential confounding in a sensitivity analysis. In EPIC-IT we dichotomized paternal profession into manual and non-manual work as a measure of occupational position. Indeed, paternal occupational position is a commonly used indicator of socioeconomic conditions in early life (Galobardes, Lynch and Smith 2007). In MESA we dichotomized highest parental education into lower if participants reported parents had no schooling or some schooling but did not complete, and higher otherwise. Parental education is another commonly used indicator of socioeconomic conditions in early life (Galobardes et al. 2007). In NICOLA a question about financial situation of the parents before age 14 was categorized into poor if participants responded poor and good if participants responded about average or pretty well off financially. In TILDA we dichotomized paternal occupational position into manual and non-manual work.

#### *Harmonization of unhealthy lifestyle and morbidities*

Self-reported smoking was classed into current, former, and never. Alcohol consumption was measured in alcohol units weekly, and we categorised participants as abstainers (0 units/week), moderate drinkers (1-21 units/week for men, 1-14 units/week for women), or harmful drinkers (>21 units/week for men, >14 units/week for women). Height and weight were measured using standard procedures; body mass index (BMI) was calculated as $kg/m^2$ and categorised as normal (<25), overweight (25 to <30), or obese (≥30). Leisure physical activity was measured with different questions in each study so we dichotomised it into low and high physical activity. Hypertension and diabetes were categorized as having or not having the morbidity.

#### *Mediation Method*

We adopted a counterfactual mediation framework to disentangle direct and indirect effects of the exposure(s) on the outcome, via one or multiple mediators. Let nested counterfactual Y(a, M(a*)) denote the survival age that would have been observed if exposure A were set to higher educational attainment (a) and mediator M (e.g. epigenetic ageing) to the value would have taken if exposure were set to lower educational attainment (a*). Denoting as S the sex of participants and g(t) the hazard function at age t, the total effect of exposure on outcome within levels of S is provided by

$$TE(a, a^{*}, s, t) = g\{E[Y(a, M(a))|S = s], t\} - g\{E[Y(a^{*}, M(a^{*}))|S = s], t\}$$

The above TE can be linearly decomposed into a pure direct (DE) and indirect (IE) (VanderWeele 2015, Tchetgen Tchetgen 2013) effect within levels of S that are defined by

$$TE = DE + IE$$

$$DE(a, a^*, s, t) = g\{E[Y(a, M(a^*))|S = s], t\} - g\{E[Y(a^*, M(a^*))|S = s], t\}$$

$$IE(a, a^*, s, t) = g\{E[Y(a, M(a))|S = s], t\} - g\{E[Y(a, M(a^*))|S = s], t\}$$

The IE represents the portion of the TE explained by the chosen mediator M, whereby exposure and mediator can interact in their effect on the outcome. In main analyses, we hypothesized that the above effects were age-invariant.

Henceforth, we will describe the IORW method implementation for estimating above effects on the absolute scale or as hazard differences. We specified the hazard function g via the semiparametric additive hazard model, introduced by McKeague and Sasieni (McKeague and Sasieni 1994) to extend the work by Aalen (Martinussen and Sheike 2006), whereby the baseline hazard was left age-variant and non-parametrized while covariates (exposure and sex in main analyses) were parametrized as linearly additive. Namely, the parametrization of g resulted in

$$g(t, A, S) = \lambda_0(t) + \lambda_1 A + \lambda_2 S + \lambda_3 AS \qquad \text{(Equ. 1)}$$

We applied the inverse odds ratio weighting (IORW) approach (Tchetgen Tchetgen 2013) to estimate DE, IE and TE given the above choice of hazard parametrization. Briefly, the total effect corresponded to the parameter $\lambda_1$ for the exposure in the above hazard model, and was estimated with the *timereg (Martinussen and Sheike 2006)* R package. The DE corresponded to the same parameter $\lambda_1$ when the model in Equ. 1 was estimated through weighted regression, whereby the chosen weights are inverse odds ratio weights considering the relationship between exposure and mediator. Indeed, inverse odds ratio weighting condenses the relationship between exposure and mediator using the odds ratio function as a measure of association into a weight, removing the necessity to specify the regression model for the outcome on exposure and mediator, including any exposure–mediator interactions (Tchetgen Tchetgen 2013). Applying the inverse odds ratio weights renders the exposure and mediator independent, deactivating the indirect pathways involving the mediator. To estimate these weights, we parametrized the relationship between exposure and mediator with the following logistic model

$$logit(A) = \beta_0 + \beta_1 M + \beta_2 S + \beta_3 MS + \beta_4 Age_{baseline} \quad \text{(Equ. 2)}$$

The weights were computed by taking the inverse of the predicted odds ratio for each observation in the exposed group. To achieve a more efficient estimation of DE, we further stabilized the weights by multiplying each individual's exposure-mediator odds ratio by the predicted odds of the exposure, where the mediator was evaluated at its reference value (Nguyen et al. 2015).

Last, the IE was estimated by taking the difference between the total effect and the direct effect on the scale used to obtain direct and total effects.

The required identifying conditions for IE, DE and TE within the IORW mediation approach are no model misspecification, no measurement errors of exposure, mediator and outcome, positivity, consistency and no unmeasured confounding for the effect of exposure on mediator, mediator on outcome possibly affected by exposure, and exposure on outcome (Tchetgen Tchetgen 2013, VanderWeele 2015). When all the above conditions are satisfied, the estimated effects can be interpreted as causal effects.

The IORW can be extended to study the mediating effect of multiple mediators *en-bloc* by simply adding the chosen mediators as predictors in the model for the exposure of Equ 2. This is parsimonious as there is no need to specify a model for the joint conditional density of mediators. Additionally, it has been shown that the identification of this joint IE does not depend upon the causal order of the mediators and upon unmeasured confounding of mediator-mediator associations (VanderWeele and Vansteelandt 2014, Steen et al. 2017).

In summary, to obtain estimates from the IORW method we (Nguyen et al. 2015): i) fitted a logistic regression model for education given mediator(s), age at baseline and sex, with an interaction between mediator(s) and sex; ii) computed stabilized weights for each participant from the predicted odds ratios at step i; iii) estimated the direct effect via a weighted additive hazard (or Cox) regression model with education, sex and their interaction as predictors; iv) estimated the total effect via an additive hazard (or Cox) regression model with education, sex and their interaction as predictors; v) estimate indirect effect as the difference between total and direct effects; vi) generated 5,000 bootstrap draws with replacement to derive standard errors for the estimands total and indirect effects.

Overall, the IORW method has three features that were relevant to our study: i) it does is not require to specify a model for the joint conditional density of multiple mediators but mediators can just be incorporated into the model of the exposure (e.g. Equ. 2); ii) it accommodates the potential interaction between exposure and mediator without the need to parametrize it; and iii) it offers a sensitivity analysis for unmeasured (possibly exposure-induced) confounding of the mediator-outcome relationship (Nguyen et al. 2015).

***Meta-analytic method***

We estimated total and indirect effects in each cohort study and then we pooled those estimates via the Hartung-Knapp inverse variance random effect meta-analytic model (Hartung and Knapp 2001). This method has been shown to provide reliable coverage accuracy in a small number study setting (Guolo and Varin 2017). However, since the Hartung-Knapp method can provide overly narrow confidence intervals when single-cohort effects are homogenous, we reported confidence intervals based on the fixed-effect model whenever that was the case as suggested by Wiksten et al. (Wiksten, Rücker and Schwarzer 2016).

The inter-study variance $\tau^2$ was estimated via the DerSimonian-Laird method implemented in the *meta (Schwarzer, Carpenter and Rücker 2015)* R package. The potential heterogeneity across studies was assessed through the $\tau^2$ and $I^2$ statistics, the former lying between 0 and 100 with higher between-study heterogeneity corresponding to higher values. We report these statistics in the forest plots at the end of the

**Supplementary Material**.

***Sensitivity analyses***

We run various sensitivity analyses to investigate the robustness of our findings with respect to assumptions of confounding, measurement error and age-invariant effects posited by our implementation of the mediation method, and to assess whether the mediation by epigenetic ageing biomarkers is related to DNA methylation levels beyond cellular heterogeneity in blood. Specifically, firstly we included childhood socioeconomic conditions as a potential confounder in a subset of four cohort studies (EPIC-IT, MESA, NICOLA, TILDA) where this information was available. This sensitivity analysis allowed us to investigate whether findings are robust with respect to this potential confounding of the exposure-outcome, exposure-mediator and mediator-outcome relationship.

Secondly, we run the sensitivity analysis proposed by Tchetgen Tchetgen and Shpitser (Tchetgen and Shpitser 2012) to assess the potential bias induced by unmeasured (possibly exposure-induced) confounding of the mediator-outcome relationship. This sensitivity analysis quantifies the potential bias in the indirect effect by offsetting the observed outcome by a value encoded in a selection bias function. We chose the selection bias function detailed in Appendix 2 of the manuscript by Nguyen et al. (Nguyen et al. 2015). This selection bias function depended on the sensitivity parameter $\rho(S)$ defined as

$$\rho(S) = E[Y(a^*, M) | M = 1, S] - E[Y(a^*, M) | M = 0, S]$$

whereby the mediator M (epigenetic ageing) was binary. The $\rho(S)$ captured the extent to which confounding may lead to differences in the average potential survival age of participants having lower educational attainment and elevated epigenetic age (epigenetic ageing > 0), compared to the average potential survival age of participants having lower educational attainment and mitigated epigenetic age (epigenetic ageing < 0). Thus, by varying $\rho(S)$ one may obtain a sensitivity analysis for confounding of the mediator. We estimated the indirect effect for values of $\rho(S)$ ranging from -5 to 5 years.

Thirdly, we assessed the impact of measuring epigenetic ageing biomarkers with error, as indirect effects can be biased in presence of an imperfectly measured mediator (VanderWeele, Valeri and Ogburn 2012). To accommodate a non-differential measurement error in the mediator, the model for the exposure in Equ. 2 implemented the SIMEX simulation-based approach for measurement error correction (Valeri, Lin and VanderWeele 2014). From a previous validation study of reproducibility in estimating epigenetic ageing biomarkers from DNA methylation in blood (McEwen et al. 2018), we hypothesized that errors were of the order of 3 years.

Fourthly, we estimated total and indirect effects by incrementally right censoring survival age at 70, 75, 80, and 85 years to assess the validity of findings based on our assumption of constant effects (Stensrud and Hernán 2020).

Lastly, we applied a sequential mediation approach (VanderWeele and Vansteelandt 2014) to assess the path-specific effect mediated by epigenetic ageing biomarkers and not leukocyte composition in blood. Specifically, we estimated the indirect effect when only leukocyte composition in blood was included as a mediator in the first step, followed by an estimation of the indirect effect when both leukocyte composition and epigenetic ageing biomarkers in blood were included as mediators. Given the posited causal structure in **Supplementary Figure 6**, a substantive additional indirect effect provided by both mediators suggests epigenetic ageing explains educational inequalities independently of leukocyte composition in blood, or in other words that there is mediation by levels of DNA methylation in blood beyond the proportion of leukocyte types in blood. We run this sensitivity analysis on all cohorts but MESA, as therein DNA methylation had been measured from monocytes.

**Supplementary Table 1**. Categorization of education.

| Cohort | Lower education | Higher education |
|---|---|---|
| **EPIC-IT** | None, primary school, lower secondary school, vocational school | Higher secondary school, BSc, MSc |
| **ESTHER** | =< 9 years (No education, or primary") | >=10-11 years ("secondary education, high school or more”) |
| **KORA** | =< 9 years (No education, or primary") | >=10-11 years ("secondary education, high school or more”) |
| **MCCS** | Never attended school, attended some primary school, completed primary school, obtained other qualifications such as a trade certificate, attended some high/technical school | Completed high school or technical school, completed a tertiary degree or diploma, completed some study towards a tertiary degree or diploma |
| **MESA** | No schooling; grades 1-8; grades 9-11 | Completed high-school/GED; some college but not degree; technical school certificate; associate degree, bachelor’s degree; graduate or professional school |
| **NICOLA** | None, some primary (not complete), Primary or equivalent, GCSE/Intermediate/junior/group certificate or equivalent, A-level/Leaving certificate or equivalent | Diploma/certificate, Primary degree, Postgraduate/higher degree |
| **RS** | Primary education; intermediate general and lower vocational education | Higher general and intermediate vocational education; higher vocational education and university |
| **TILDA** | None, some primary (not complete), Primary or equivalent, Intermediate/junior/group certificate or equivalent, Leaving certificate or equivalent | Diploma/certificate, Primary degree, Postgraduate/higher degree |

**Supplementary Table 2**. Country, birth year and time-line for collection of exposure and mediator data in each cohort. The reported range of birth years corresponds to the 10th and 90th quantiles of the distribution.

| Cohort | Country | Birth year | Exposure | Mediators |
|---|---|---|---|---|
| **EPIC-IT** | Italy | 1933 – 1952 | 1993 –1998 | 1993 – 1998 |
| **ESTHER** | Germany | 1929 – 1948 | 2000 – 2001 | 2000 – 2001 |
| **KORA** | Germany | 1934 – 1958 | 1999 – 2001 | 2006 – 2008 |
| **MCCS** | Australia | 1913 – 1936 | 1990 – 1994 | 1990 – 1994 |
| **MESA** | USA | 1929 – 1954 | 2000 – 2002 | 2010 – 2012 |
| **NICOLA** | Northern Ireland | 1937 – 1962 | 2014 – 2016 | 2014 – 2016 |
| **RS** | The Netherlands | 1937 – 1956 | 2007 – 2012 | 2007 – 2012 |
| **TILDA** | Ireland | 1936 – 1958 | 2009 – 2011 | 2009 – 2011 |

**Supplementary Table 3. Mediation by unhealthy lifestyle, morbidities, and epigenetic ageing in women**. Absolute (hazard difference per 10,000 person-years) and relative (hazard ratio) size of total effect of lower (vs higher) educational attainment on all-cause mortality, and of indirect effects by unhealthy lifestyle and morbidities (LM) and epigenetic ageing biomarkers (Horvath DNAmAA, Hannum DNAmAA, DNAmPhenoAA, and DNAmGrimAA). All effects are pooled estimates of single cohort's effects through a weighted inverse variance meta-analytic model. The total number of participants/deaths across cohorts is 6,175 / 964.

| EFFECTS | Hazard difference (95% CI) | Hazard ratio (95% CI) |
|---|---|---|
| Total effect | 6 (-14 to 26) | 1.17 (1.05 to 1.30) |
| Indirect effect by LM | -5 (-12 to 2) | 1.02 (0.94 to 1.11) |
| Indirect effect by LM and Horvath DNAmAA | -4 (-10 to 2) | 1.03 (0.98 to 1.02) |
| Indirect effect by LM and Hannum DNAmAA | -5 (-11 to 2) | 1.02 (0.93 to 1.12) |
| Indirect effect by LM and DNAmPhenoAA | -5 (-11 to 1) | 1.02 (0.95 to 1.09) |
| Indirect effect by LM and DNAmGrimAA | -3 (-11 to 4) | 1.03 (0.95 to 1.12) |

**Supplementary Table 4. Potential confounding by childhood socioeconomic conditions.** Absolute effects quantifying total effect of education (lower vs higher) on all-cause mortality and indirect effect by epigenetic ageing biomarkers (Horvath DNAmAA, Hannum DNAmAA, DNAmPhenoAA, and DNAmGrimAA) when childhood socioeconomic conditions (CSC) were included as an additional potential confounding. All effects are pooled estimates of single cohort's hazard ratios through a weighted inverse variance meta-analytic model. The total number of participants/deaths across EPIC-IT, MESA, NICOLA, and TILDA cohorts is 2,370 / 290 for men and 2,674 / 225 for women.

| **EFFECTS** | **Men** | | **Women** | |
|---|---|---|---|---|
| | **Without CSC** | **With CSC** | **Without CSC** | **With CSC** |
| Total effect | 56 (32 to 79) | 57 (28 to 85) | 6 (-11 to 23) | 6 (-11 to 24) |
| Indirect effect by Horvath DNAmAA | 12 (4 to 20) | 13 (8 to 17) | -5 (-8 to -1) | -4 (-7 to -1) |
| Indirect effect by Hannum DNAmAA | 17 (9 to 25) | 19 (12 to 25) | -5 (-9 to -1) | -4 (-9 to 0) |
| Indirect effect by DNAmPhenoAA | 19 (9 to 30) | 21 (14 to 27) | -5 (-8 to -2) | -5 (-8 to -2) |
| Indirect effect by DNAmGrimAA | 29 (21 to 38) | 30 (22 to 38) | -1 (-15 to 13) | 1 (-13 to 14) |

**Supplementary Table 5. Path-specific indirect effect by epigenetic ageing independent of leukocyte composition in men**. Absolute (hazard difference per 10,000 person-years) size of total effect of lower (vs higher) educational attainment on all-cause mortality, and for indirect effects by leukocyte composition (LC) and epigenetic ageing in blood. Four epigenetic ageing biomarkers based on different epigenetic clocks estimators are reported: Horvath DNAmAA, Hannum DNAmAA, DNAmPhenoAA, and DNAmGrimAA. All effects are pooled estimates of single cohort's hazards through a weighted inverse variance meta-analytic model. The study MESA is not included as therein epigenetic ageing biomarkers were computed from DNA methylation in monocytes. The total number of participants/deaths across cohorts is 5,865 / 1,559.

| EFFECTS | Hazard difference (95% CI) |
|---|---|
| Total effect | 57 (39 to 74) |
| Indirect effect by LC | 16 (10 to 22) |
| Indirect effect by LC and Horvath DNAmAA | 15 (8 to 22) |
| Indirect effect by Horvath DNAmAA but not LC | 0 (-1 to 1) |
| Indirect effect by LC and Hannum DNAmAA | 18 (12 to 24) |
| Indirect effect by Hannum DNAmAA but not LC | 2 (0 to 4) |
| Indirect effect by LC and DNAmPhenoAA | 21 (14 to 27) |
| Indirect effect by DNAmPhenoAA but not LC | 5 (2 to 7) |
| Indirect effect by LC and DNAmGrimAA | 34 (26 to 43) |
| Indirect effect by DNAmGrimAA and not LC | 18 (10 to 25) |

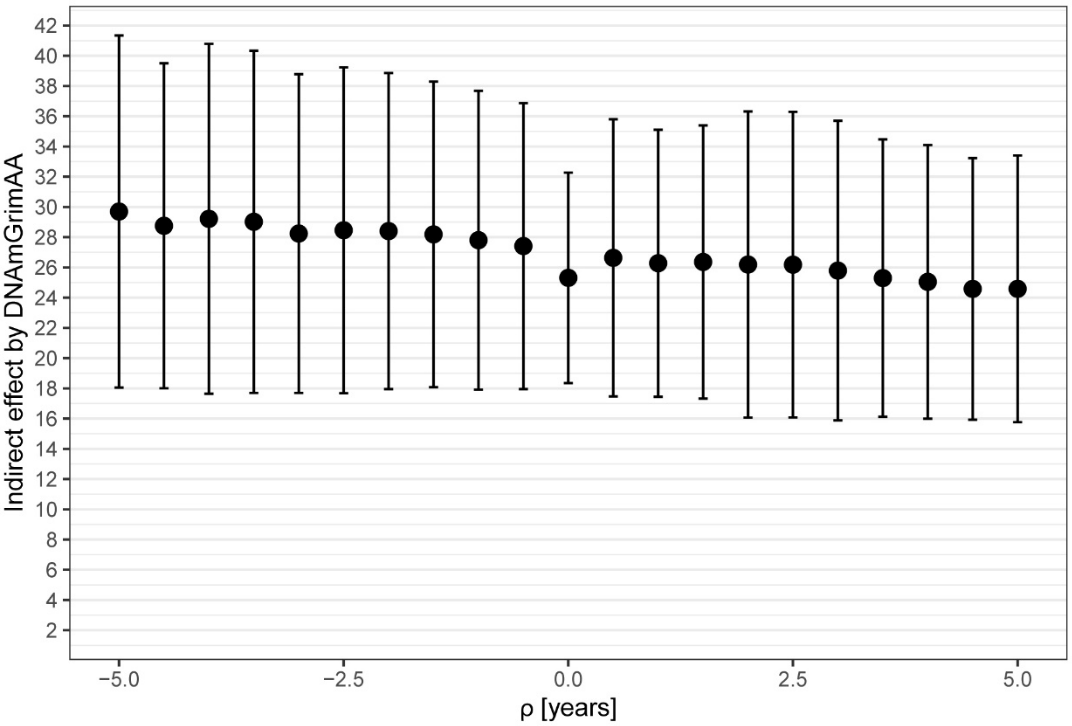


**Supplementary Figure 1**. **Indirect effect by epigenetic ageing biomarker DNAmGrimAA in men as a function of bias (ρ) in the outcome due to unmeasured confounding of the mediator-outcome relationship.** Rho represents the counterfactual difference in average survival age for being exposed to lower education and elevated epigenetic age (positive values of DNAmGrimAA) compared to being exposed to lower education and mitigated epigenetic age (negative values of DNAmGrimAA). A positive value of ρ indicates a delayed survival age due to a hypothetical intervention for slowing or mitigating epigenetic ageing, while a negative value indicates the opposite. Pooled excess deaths per 10,000 person-years and 95% compatibility intervals (CI) are reported. All effects are pooled estimates of single cohort's hazard differences through a weighted inverse variance meta-analytic model. The total number of participants/deaths across cohorts is 6,477 / 1,638.

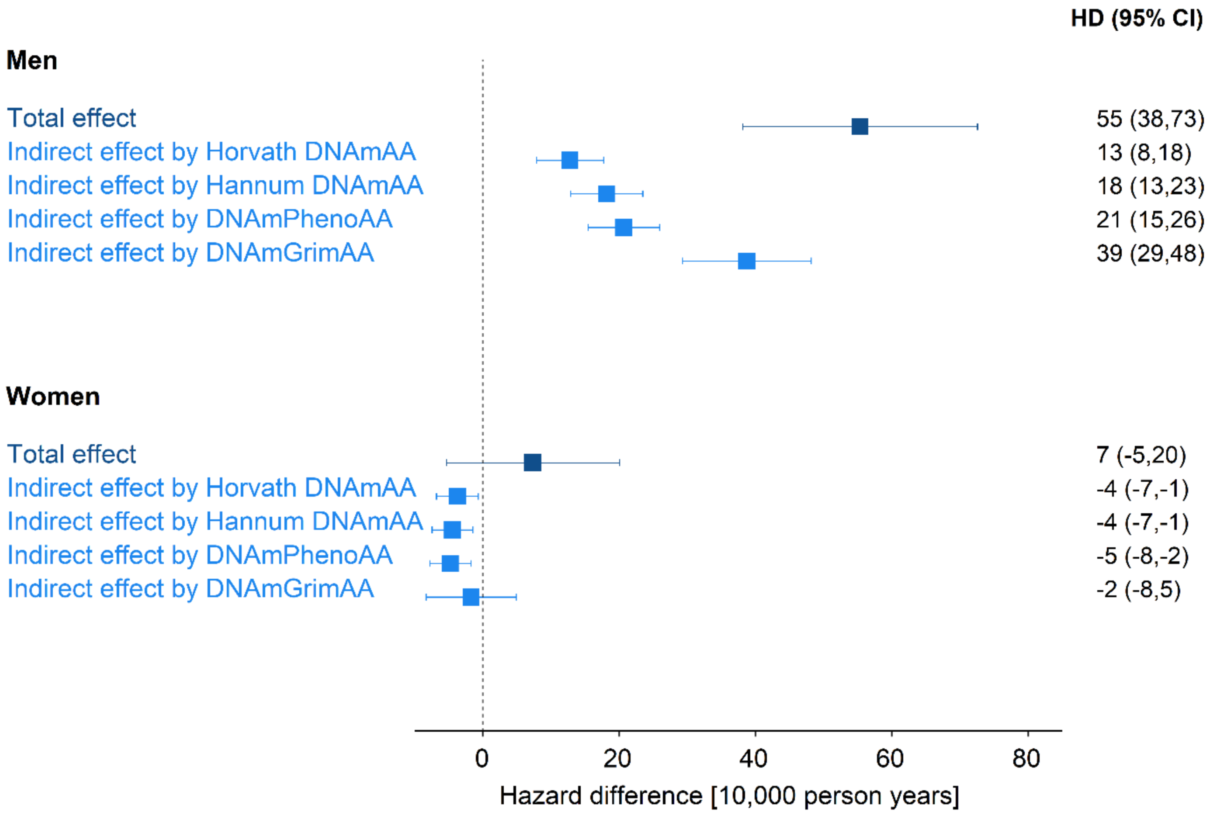


**Supplementary Figure 2. Educational inequalities and mediation by epigenetic ageing biomarkers on the absolute scale when considering a potential measurement error of three years in the assessment of the biomarkers**. Excess deaths per 10,000 person-years and 95% compatibility intervals (CI) are reported for the total effect of lower (vs higher) educational attainment on all-cause mortality, and for the indirect effect by epigenetic ageing biomarkers (Horvath DNAmAA, Hannum DNAmAA, DNAmPhenoAA, and DNAmGrimAA). All effects are pooled estimates of single cohort's hazard differences through a weighted inverse variance meta-analytic model. The total number of participants/deaths across cohorts is 6,477 / 1,638 for men and 6,544 / 1,002 for women.

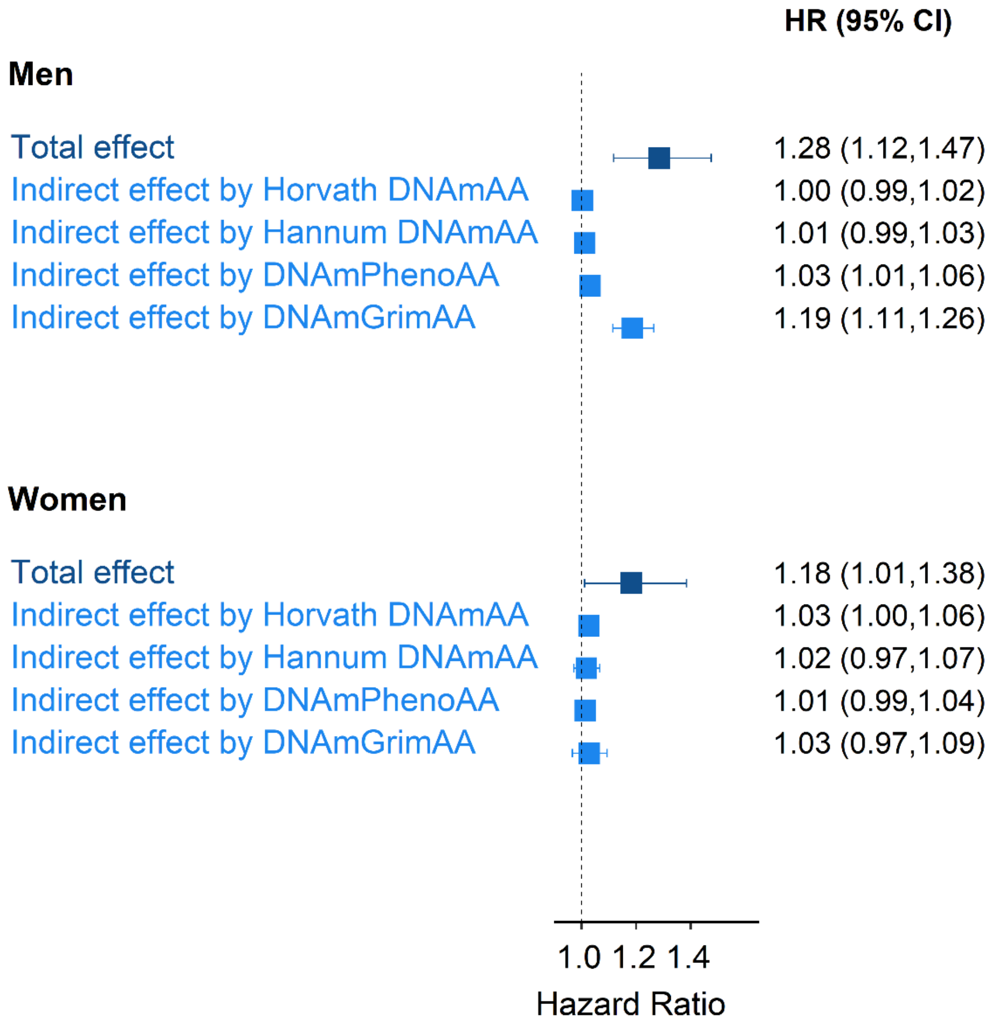


**Supplementary Figure 3**. **Educational inequalities and mediation by epigenetic ageing biomarkers on the relative scale when considering a potential measurement error of three years in the assessment of the biomarkers**. Hazard ratios and 95% compatibility intervals (CI) are reported for the total effect of lower (vs higher) educational attainment on all-cause mortality, and for the indirect effect by epigenetic ageing biomarkers (Horvath DNAmAA, Hannum DNAmAA, DNAmPhenoAA, and DNAmGrimAA). All effects are pooled estimates of single cohort's hazard ratios through a weighted inverse variance meta-analytic model. The total number of participants/deaths across cohorts is 6,477 / 1,638 for men and 6,544 / 1,002 for women.

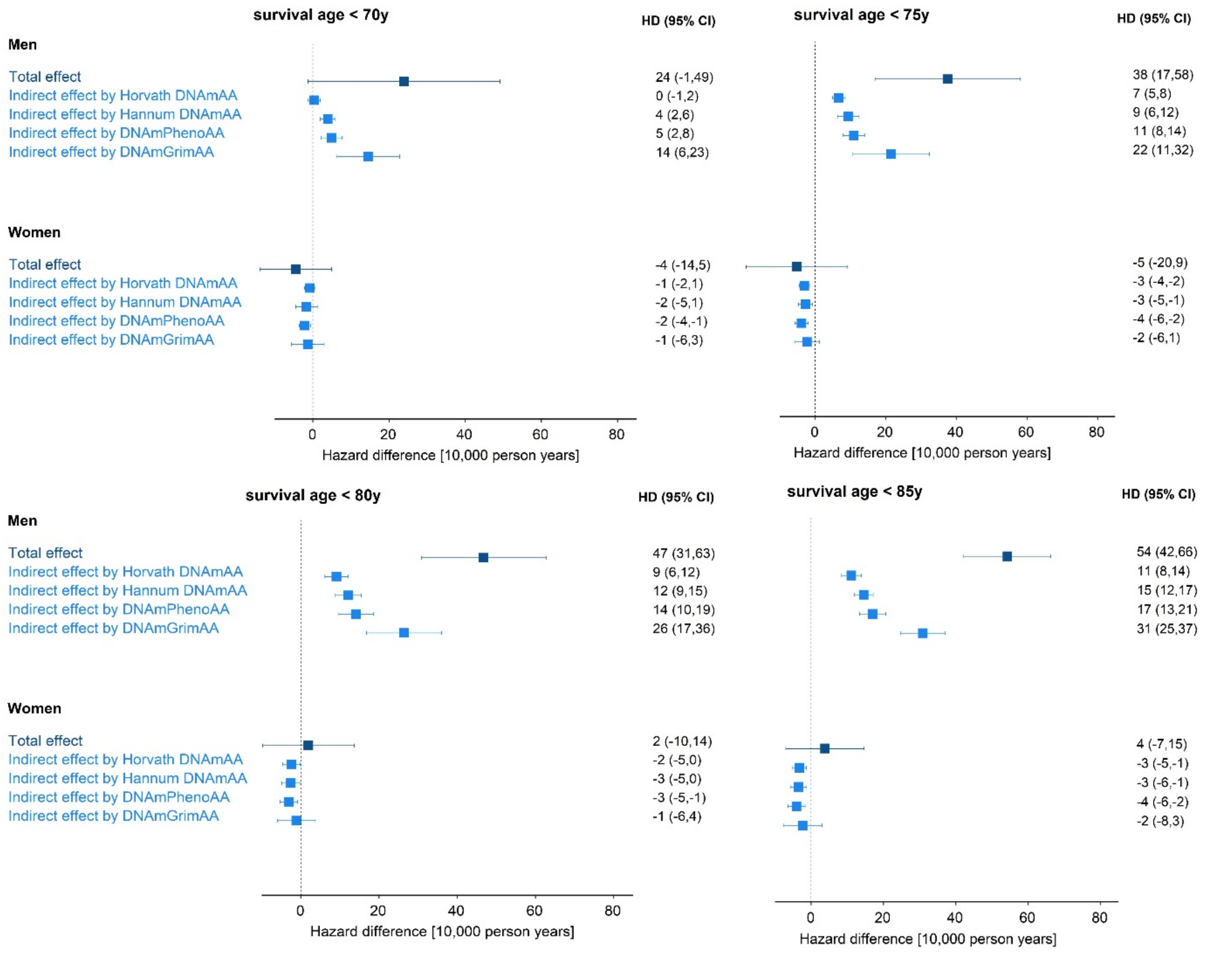


**Supplementary Figure 4**. **Educational inequalities and mediation by epigenetic ageing biomarkers on the absolute scale at incremental right censoring values of survival age**. Hazard difference per 10,000 person-years and 95% compatibility intervals (CI) are reported for the total effect of lower (vs higher) educational attainment on all-cause mortality, and for indirect effect by epigenetic ageing biomarkers (Horvath DNAmAA, Hannum DNAmAA, DNAmPhenoAA, and DNAmGrimAA) when survival age is right censored at 70y, 75y, 80y, and 85y (upper left, upper right, lower left, lower right panels, respectively). All effects are pooled estimates of single cohort's hazard differences through a weighted inverse variance meta-analytic model. The total number of participants/deaths across cohorts is 6,477 / 1,638 for men and 6,544 / 1,002 for women.

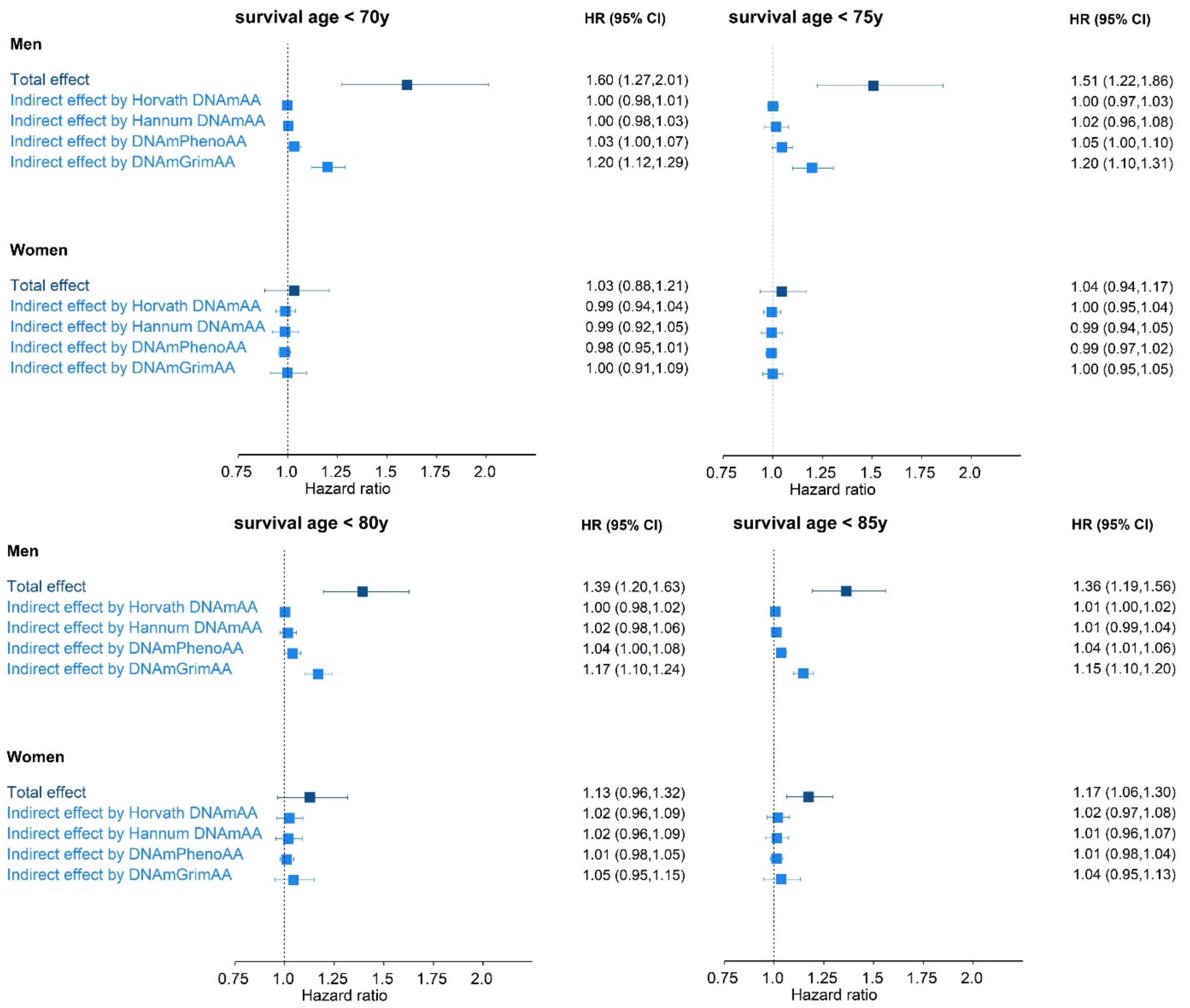


**Supplementary Figure 5**. **Educational inequalities and mediation by epigenetic ageing biomarkers on the relative scale at incremental right censoring values of survival age**. Hazard ratios and 95% compatibility intervals (CI) are reported for the total effect of lower (vs higher) educational attainment on all-cause mortality, and for indirect effect by epigenetic ageing biomarkers (Horvath DNAmAA, Hannum DNAmAA, DNAmPhenoAA, and DNAmGrimAA) when survival age is right censored at 70y, 75y, 80y, and 85y (upper left, upper right, lower left, lower right panels, respectively). All effects are pooled estimates of single cohort’s hazard ratios through a weighted inverse variance meta-analytic model. The total number of participants/deaths across cohorts is 6,477 / 1,638 for men and 6,544 / 1,002 for women.

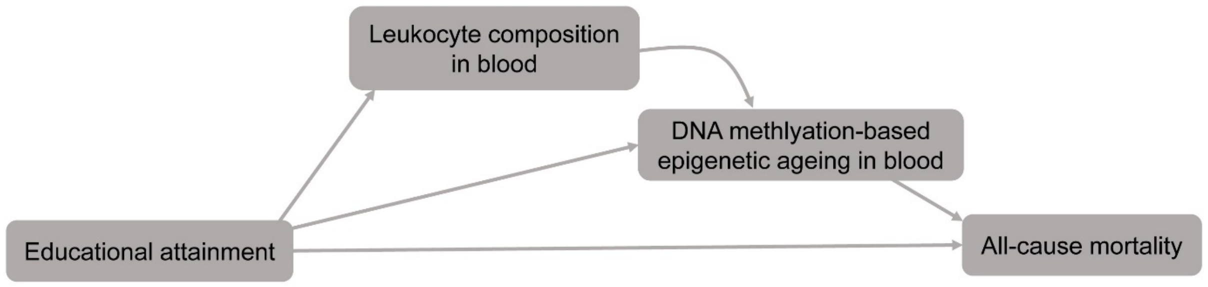


**Supplementary Figure 6. Causal structure to assess the path-specific effect of epigenetic ageing biomarkers independent of leukocyte composition in blood.** For the sake of simplicity, we do not draw confounders (age and sex). Educational attainment represents the exposure and all-cause mortality the outcome. Leukocyte composition in blood and DNA methylation-based epigenetic ageing in blood are the mediators.

***Forest Plots***

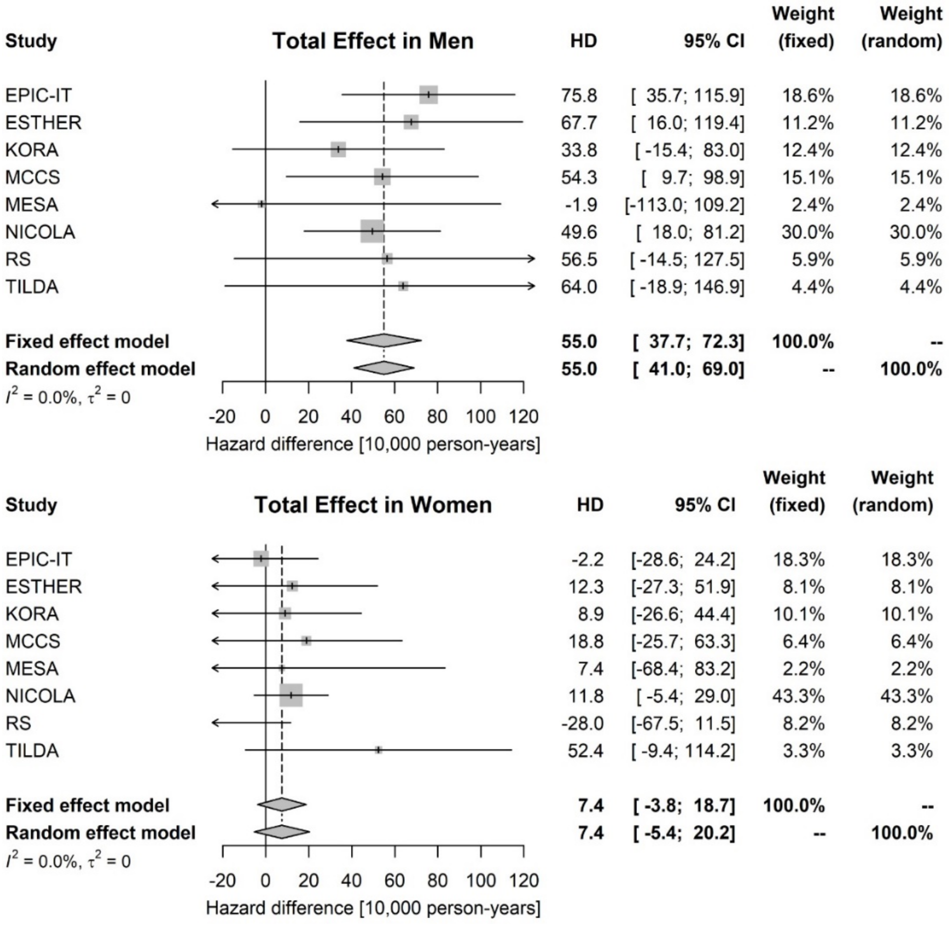


**Supplementary Figure 7. Forest plots of the total effect of educational attainment on all-cause mortality in men (top) and women (bottom) on the absolute scale.** Hazard difference (HD) per 10,000 person-years and 95% compatibility intervals (CI) are reported. Weight represents the contribution of each study to the pooled estimates (diamond) as computed by the fixed/random meta-analytic effect model. Heterogeneity across studies is quantified by $I^2$ and $\tau^2$.

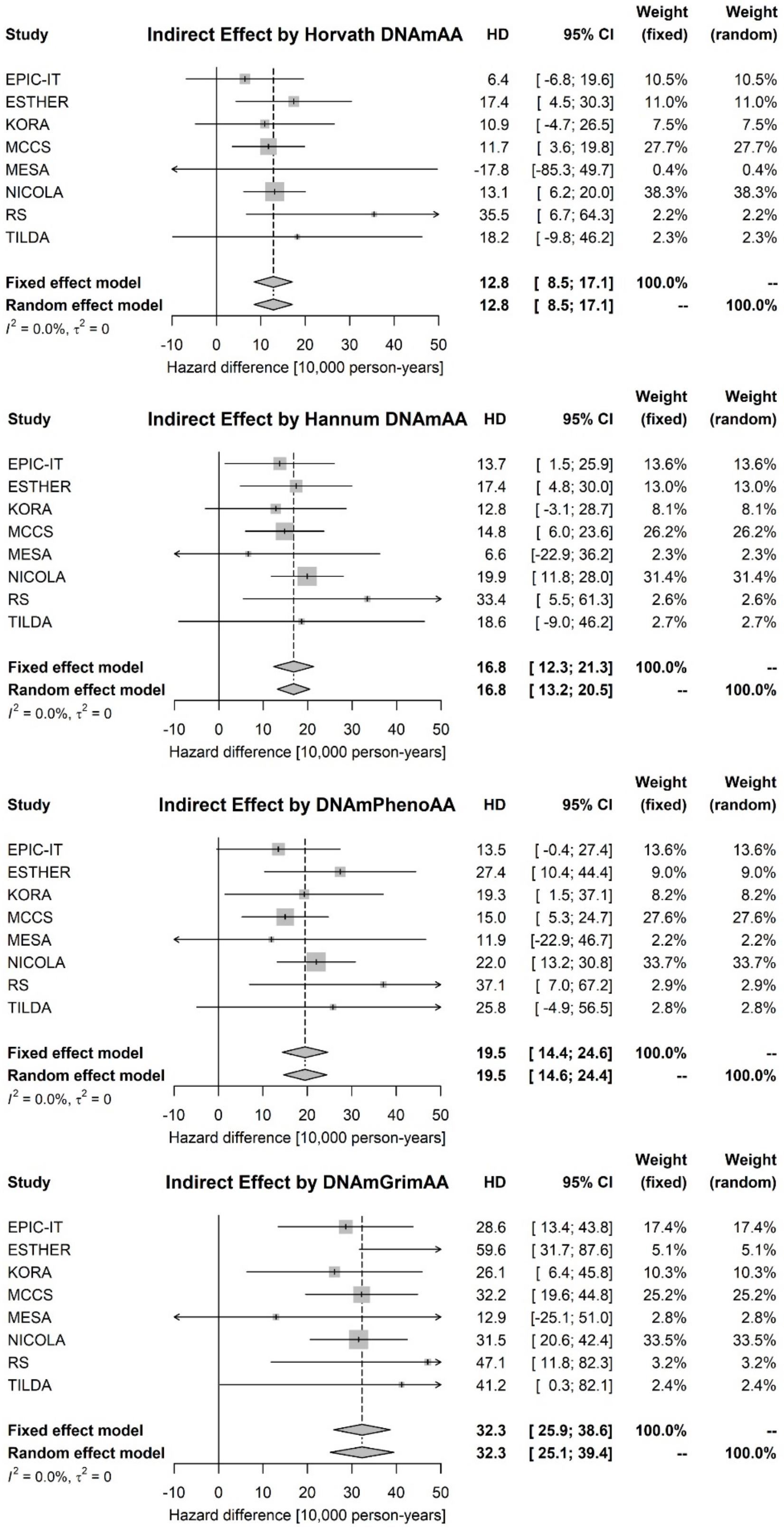


**Supplementary Figure 8. Forest plots of the indirect effect by epigenetic ageing biomarkers in men on the absolute scale.** Hazard difference (HD) per 10,000 person-years and 95% compatibility intervals (CI) are reported. Weight represents the contribution of each study to the pooled estimates (diamond) as computed by the fixed/random meta-analytic effect model. Heterogeneity across studies is quantified by $I^2$ and $\tau^2$.

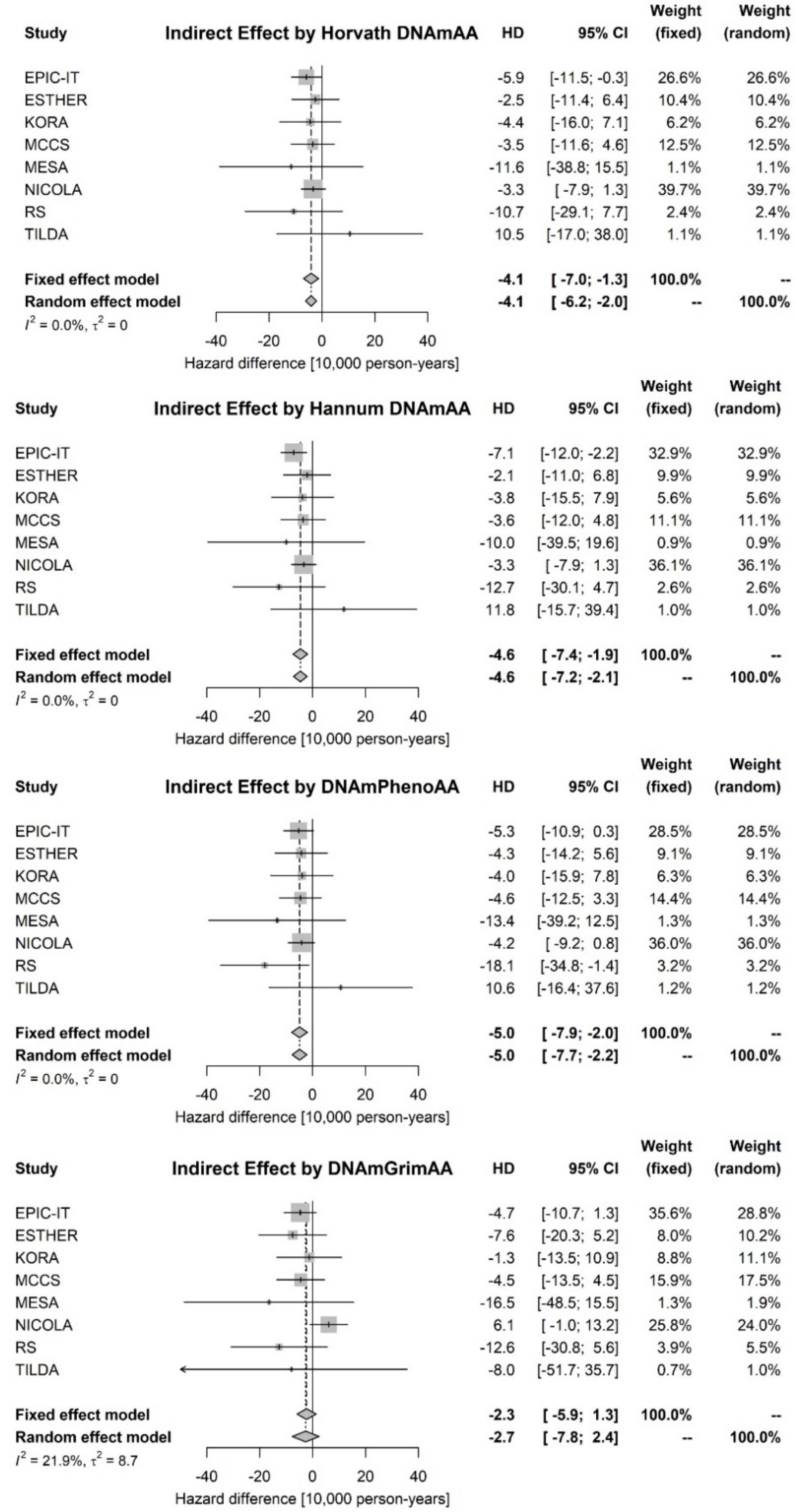


**Supplementary Figure 9. Forest plots of the absolute indirect effect by epigenetic ageing biomarkers in women on the absolute scale.** Hazard difference (HD) per 10,000 person-years and 95% compatibility intervals (CI) are reported. Weight represents the contribution of each study to the pooled estimates (diamond) as computed by the fixed/random meta-analytic effect model. Heterogeneity across studies is quantified by $I^2$ and $\tau^2$.

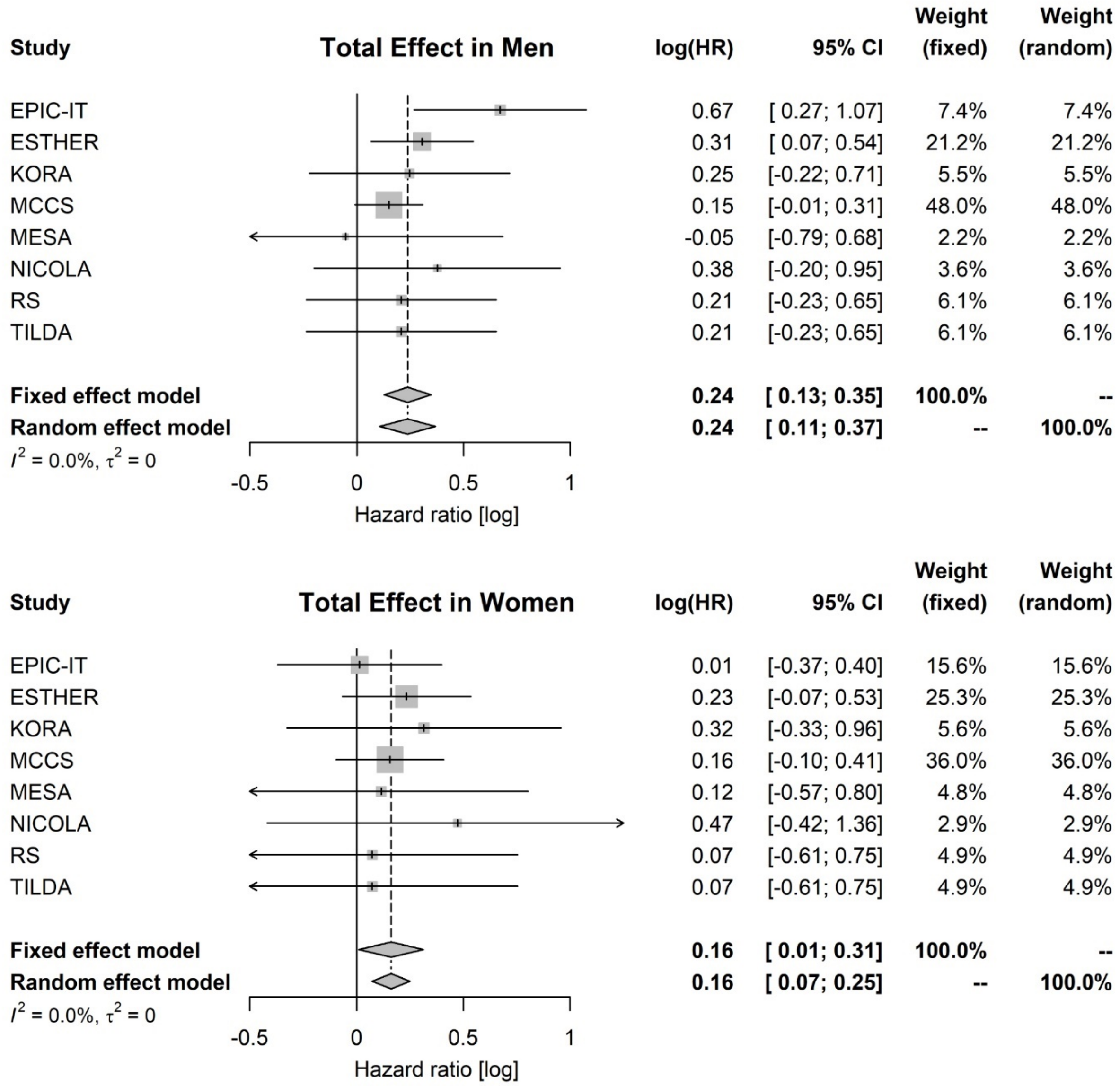


**Supplementary Figure 10. Forest plots of the relative effect of educational attainment on all-cause mortality in men (top) and women (bottom) on the relative scale.** Hazard ratio (HR) and 95% compatibility intervals (CI) are reported. Weight represents the contribution of each study to the pooled estimates (diamond) as computed by the fixed/random meta-analytic effect model. Heterogeneity across studies is quantified by $I^2$ and $\tau^2$.

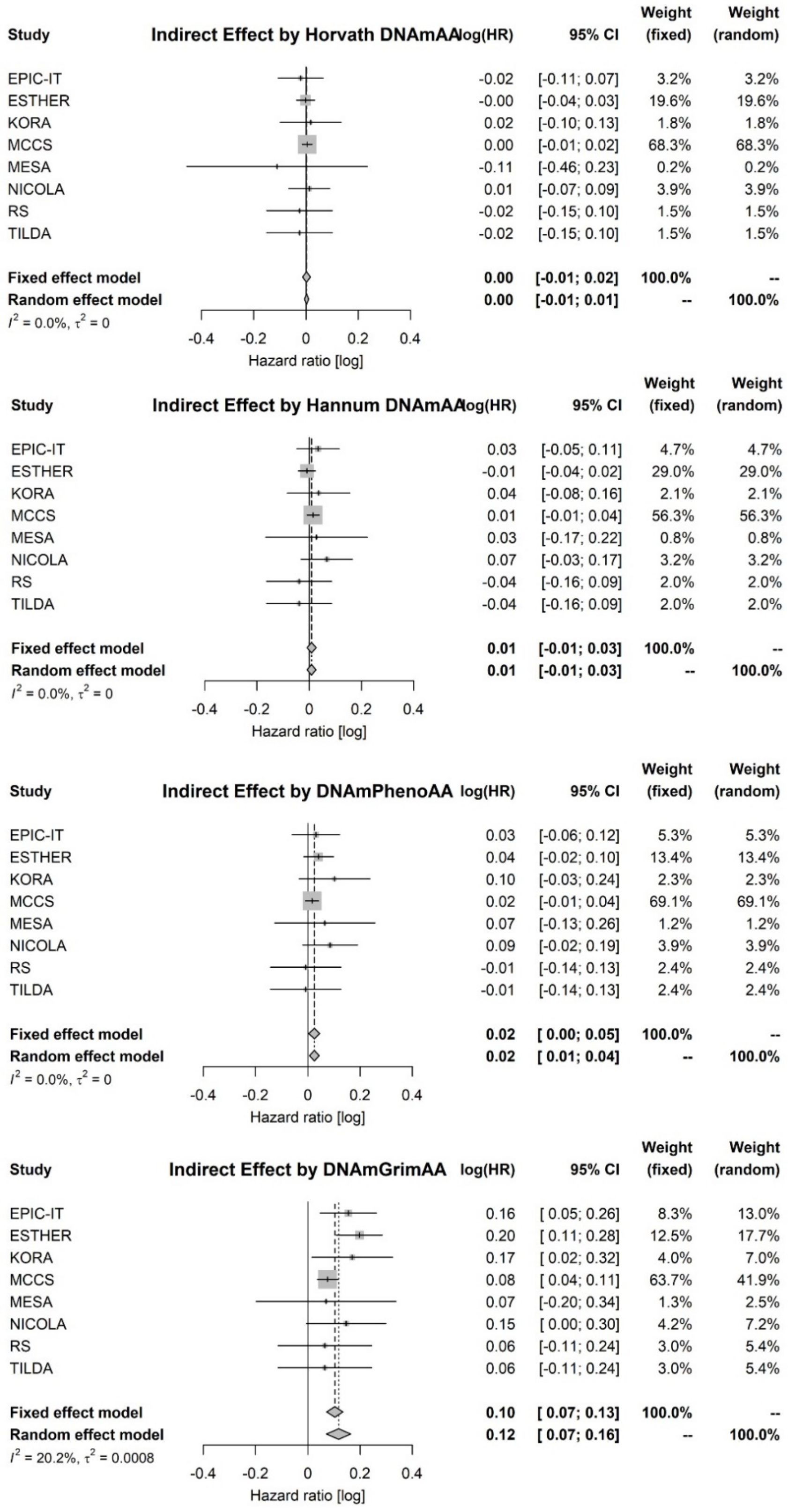


**Supplementary Figure 11. Forest plots of the relative indirect effect by epigenetic ageing biomarkers in men on the relative scale.** Hazard ratio (HR) and 95% compatibility intervals (CI) are reported. Weight represents the contribution of each study to the pooled estimates (diamond) as computed by the fixed/random meta-analytic effect model. Heterogeneity across studies is quantified by $I^2$ and $\tau^2$.

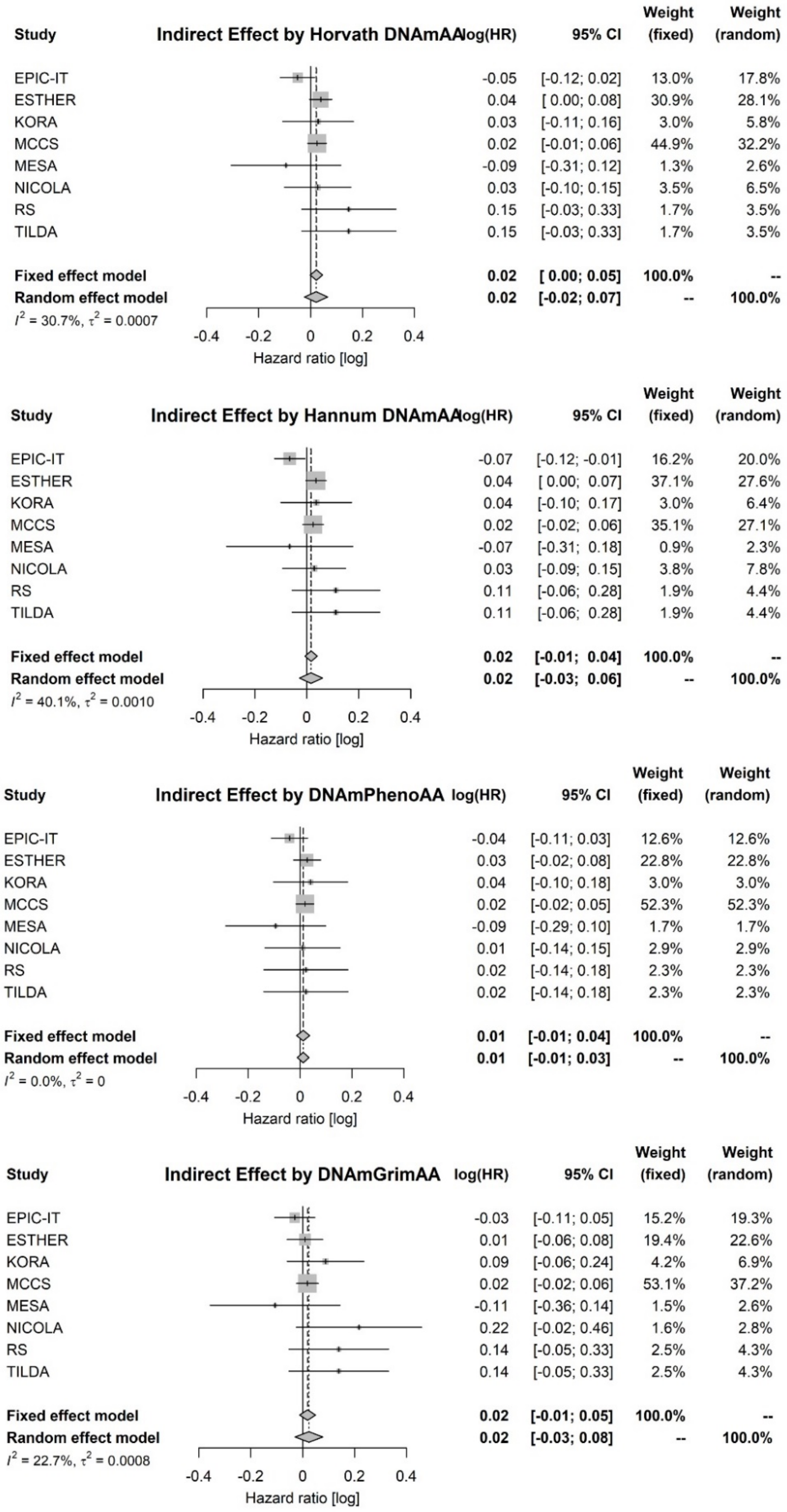


**Supplementary Figure 12. Forest plots of the relative indirect effect by epigenetic ageing biomarkers in women on the relative scale.** Hazard ratio (HR) and 95% compatibility intervals (CI) are reported. Weight represents the contribution of each study to the pooled estimates (diamond) as computed by the fixed/random meta-analytic effect model. Heterogeneity across studies is quantified by $I^2$ and $\tau^2$.

### *Extended acknowledgements*


| Cohort | |
|---|---|
| **EPIC-IT** | EPIC-Italy was funded by the Associazione Italiana Ricerca contro il Cancro (AIRC). |
| **ESTHER** | The ESTHER study was funded by the Baden-Württemberg State Ministry of Science, Research and Arts (Stuttgart, Germany), the German Federal Ministry of Education and Research (Berlin, Germany), the German Federal Ministry of Family, Senior Citizens, Women and Youth (Berlin, Germany) and the Saarland State Ministry of Social Affairs, Health, Women and Family (Saarbrücken, Germany). The analyses for this project were supported by a grant from the German Cancer Aid (No. 70113330). |
| **KORA** | The KORA study was initiated and financed by the Helmholtz Zentrum München – German Research Center for Environmental Health, which is funded by the German Federal Ministry of Education and Research (BMBF) and by the State of Bavaria. Furthermore, KORA research has been supported within the Munich Center of Health Sciences (MC-Health), Ludwig-Maximilians-Universität, as part of LMUinnovativ. |
| **MCCS** | Melbourne Collaborative Cohort Study (MCCS) cohort recruitment was funded by VicHealth and Cancer Council Victoria. The MCCS was further augmented by Australian National Health and Medical Research Council grants 209057, 396414 and 1074383 and by infrastructure provided by Cancer Council Victoria. Cases and their vital status were ascertained through the Victorian Cancer Registry and the Australian Institute of Health and Welfare, including the National Death Index and the Australian Cancer Database. |
| **MESA** | MESA and the MESA SHARe project are conducted and supported by the National Heart, Lung, and Blood Institute (NHLBI) in collaboration with MESA investigators. Support for MESA is provided by contracts 75N92020D00001, HHSN268201500003I, N01-HC-95159, 75N92020D00005, N01-HC-95160, 75N92020D00002, N01-HC-95161, 75N92020D00003, N01-HC-95162, 75N92020D00006, N01-HC-95163, 75N92020D00004, N01-HC-95164, 75N92020D00007, N01-HC-95165, N01-HC-95166, N01-HC-95167, N01-HC-95168, N01-HC-95169, UL1-TR-000040, UL1-TR-001079, UL1-TR-001420, UL1-TR-001881, and DK063491. The MESA Epigenomics & Transcriptomics Studies were funded by NIH grants 1R01HL101250, 1RF1AG054474, R01HL126477, R01DK101921, and R01HL135009. Analysis was partially supported by R01HL141292.<br>This manuscript has been reviewed by MESA for scientific content and consistency of data interpretation with previous MESA publications. |
| **NICOLA** | We are grateful to all the participants of the NICOLA Study, and the whole NICOLA team, which includes nursing staff, research scientists, clerical staff, computer and laboratory technicians, managers and receptionists. The Atlantic Philanthropies, the Economic and Social Research Council, the UKCRC Centre of Excellence for Public Health Northern Ireland, the Centre for |

| | |
|---|---|
| | Ageing Research and Development in Ireland, the Office of the First Minister and Deputy First Minister, the Health and Social Care Research and Development Division of the Public Health Agency, the Wellcome Trust/Wolfson Foundation and Queen's University Belfast provide core financial support for NICOLA. The authors alone are responsible for the interpretation of the data and any views or opinions presented are solely those of the authors and do not necessarily represent those of the NICOLA Study team. |
| **RS** | The Rotterdam Study (RS) is supported by grants from the Municipality of Rotterdam, the NESTOR program for research in the elderly (Ministry of Health and Ministry of Education), the Netherlands Prevention Fund, the Netherlands Organization for scientific research (NWO), the Netherlands Heart Foundation and the Rotterdam Medical Research Foundation (ROMERES). |
| **TILDA** | Funding for the TILDA project was supported by the Irish Government, the Atlantic Philanthropies, and Irish Life plc. |

***Ethical oversight***

| Cohort name | Ethical Review Board / Institutional Review Board | Decision |
|---|---|---|
| EPIC Italy | Ethical review board of the International Agency for Research on Cancer (IARC) | Approved |
| ESTHER | Ethics committees of the University of Heidelberg and of the Medical Association of Saarland | Approved |
| MCCS | The Cancer Council Victoria's Human Research Ethics Committee | Approved |
| KORA | The Bavarian Chamber of Physicians Ethics Board | Approved |
| MESA | Institutional review boards (IRB) at each participating centre and Johns Hopkins School of Medicine IRB | Approved |
| NICOLA | the School of Medicine, Dentistry and Biomedical Sciences Ethics Committee, Queen’s University Belfast | Approved |
| Rotterdam Study | Medical ethics committee of the Erasmus University Medical Centre, Rotterdam | Approved |
| TILDA | Faculty of Health Sciences Research Ethics Committee in Trinity College, Dublin | Approved |

Each study was performed in accordance with the Declaration of Helsinki, including written informed consent from all participants.

### References


Aryee, M. J., A. E. Jaffe, H. Corrada-Bravo, C. Ladd-Acosta, A. P. Feinberg, K. D. Hansen & R. A. Irizarry (2014) Minfi: a flexible and comprehensive Bioconductor package for the analysis of Infinium DNA methylation microarrays. *Bioinformatics,* 30, 1363-9.

Baglietto, L., E. Ponzi, P. Haycock, A. Hodge, M. Bianca Assumma, C. H. Jung, J. Chung, F. Fasanelli, F. Guida, G. Campanella, M. Chadeau-Hyam, K. Grankvist, M. Johansson, U. Ala, P. Provero, E. M. Wong, J. Joo, D. R. English, N. Kazmi, E. Lund, C. Faltus, R. Kaaks, A. Risch, M. Barrdahl, T. M. Sandanger, M. C. Southey, G. G. Giles, P. Vineis, S. Polidoro, C. L. Relton & G. Severi (2017) DNA methylation changes measured in pre-diagnostic peripheral blood samples are associated with smoking and lung cancer risk. *Int J Cancer,* 140, 50-61.

Burns, F, G. Carney, S. Cruise, P. Devine, A. Devlin, M. Donnelly, D. French, F. Kee, L. Montgomery, D. O'Reilly, A. Scott & M. Tully. 2017. Early key findings from a study of older people in Northern Ireland. The NICOLA Study. Belfast: Queen's University, Belfast.

Campanella, G., S. Polidoro, C. Di Gaetano, G. Fiorito, S. Guarrera, V. Krogh, D. Palli, S. Panico, C. Sacerdote, R. Tumino, P. Elliott, G. Matullo, M. Chadeau-Hyam & P. Vineis (2015) Epigenetic signatures of internal migration in Italy. *Int J Epidemiol,* 44, 1442-1449.

Du, P., W. A. Kibbe & S. M. Lin (2008) lumi: a pipeline for processing Illumina microarray. *Bioinformatics,* 24, 1547-8.

Dugué, P. A., J. K. Bassett, J. E. Joo, L. Baglietto, C. H. Jung, E. M. Wong, G. Fiorito, D. Schmidt, E. Makalic, S. Li, M. Moreno-Betancur, D. D. Buchanan, P. Vineis, D. R. English, J. L. Hopper, G. Severi, M. C. Southey, G. G. Giles & R. L. Milne (2018) Association of DNA Methylation-Based Biological Age With Health Risk Factors and Overall and Cause-Specific Mortality. *Am J Epidemiol,* 187, 529-538.

Dugué, P. A., M. T. Brinkman, R. L. Milne, E. M. Wong, L. M. FitzGerald, J. K. Bassett, J. E. Joo, C. H. Jung, E. Makalic, D. F. Schmidt, D. J. Park, J. Chung, A. D. Ta, D. M. Bolton, A. Lonie, A. Longano, J. L. Hopper, G. Severi, R. Saffery, D. R. English, M. C. Southey & G. G. Giles (2016) Genome-wide measures of DNA methylation in peripheral blood and the risk of urothelial cell carcinoma: a prospective nested case-control study. *Br J Cancer,* 115, 664-73.

Galobardes, B., J. Lynch & G. D. Smith (2007) Measuring socioeconomic position in health research. *Br Med Bull,* 81-82, 21-37.

Guolo, A. & C. Varin (2017) Random-effects meta-analysis: the number of studies matters. *Stat Methods Med Res,* 26, 1500-1518.

Hartung, J. & G. Knapp (2001) On tests of the overall treatment effect in meta-analysis with normally distributed responses. *Stat Med,* 20, 1771-82.

Holle, R., M. Happich, H. Löwel, H. E. Wichmann & M. K. S. Group (2005) KORA--a research platform for population based health research. *Gesundheitswesen,* 67 Suppl 1, S19-25.

Houseman, E. A., W. P. Accomando, D. C. Koestler, B. C. Christensen, C. J. Marsit, H. H. Nelson, J. K. Wiencke & K. T. Kelsey (2012) DNA methylation arrays as surrogate measures of cell mixture distribution. *BMC Bioinformatics,* 13, 86.

Leek, J. T., W. E. Johnson, H. S. Parker, A. E. Jaffe & J. D. Storey (2012) The sva package for removing batch effects and other unwanted variation in high-throughput experiments. *Bioinformatics,* 28, 882-3.

Lehne, B., A. W. Drong, M. Loh, W. Zhang, W. R. Scott, S. T. Tan, U. Afzal, J. Scott, M. R. Jarvelin, P. Elliott, M. I. McCarthy, J. S. Kooner & J. C. Chambers (2015) A coherent approach for analysis of the Illumina HumanMethylation450 BeadChip improves data quality and performance in epigenome-wide association studies. *Genome Biol,* 16, 37.

Liu, Y., J. Ding, L. M. Reynolds, K. Lohman, T. C. Register, A. De La Fuente, T. D. Howard, G. A. Hawkins, W. Cui, J. Morris, S. G. Smith, R. G. Barr, J. D. Kaufman, G. L. Burke, W. Post, S. Shea, C. E. McCall, D. Siscovick, D. R. Jacobs, R. P. Tracy, D. M. Herrington & I. Hoeschele (2013) Methylomics of gene expression in human monocytes. *Hum Mol Genet,* 22, 5065-74.

Martinussen, T. & T. H. Sheike. 2006. *Dynamic regression models for survival data*. New York: Springer.

McEwen, L. M., M. J. Jones, D. T. S. Lin, R. D. Edgar, L. T. Husquin, J. L. MacIsaac, K. E. Ramadori, A. M. Morin, C. F. Rider, C. Carlsten, L. Quintana-Murci, S. Horvath & M. S. Kobor (2018) Systematic evaluation of DNA methylation age estimation with common preprocessing methods and the Infinium MethylationEPIC BeadChip array. *Clin Epigenetics,* 10**,** 123.

McKeague, I. W. & P. D. Sasieni (1994) A partly parametric additive risk model. *Biometrika,* 81**,** 501-514.

Milne, R. L., A. S. Fletcher, R. J. MacInnis, A. M. Hodge, A. H. Hopkins, J. K. Bassett, F. J. Bruinsma, B. M. Lynch, P. A. Dugué, H. Jayasekara, M. T. Brinkman, L. V. Popowski, L. Baglietto, G. Severi, K. O'Dea, J. L. Hopper, M. C. Southey, D. R. English & G. G. Giles (2017) Cohort Profile: The Melbourne Collaborative Cohort Study (Health 2020). *Int J Epidemiol,* 46**,** 1757-1757i.

Nguyen, Q. C., T. L. Osypuk, N. M. Schmidt, M. M. Glymour & E. J. Tchetgen Tchetgen (2015) Practical guidance for conducting mediation analysis with multiple mediators using inverse odds ratio weighting. *Am J Epidemiol,* 181**,** 349-56.

Palli, D., F. Berrino, P. Vineis, R. Tumino, S. Panico, G. Masala, C. Saieva, S. Salvini, M. Ceroti, V. Pala, S. Sieri, G. Frasca, M. C. Giurdanella, C. Sacerdote, L. Fiorini, E. Celentano, R. Galasso, A. Decarli, V. Krogh & EPIC-Italy (2003) A molecular epidemiology project on diet and cancer: the EPIC-Italy Prospective Study. Design and baseline characteristics of participants. *Tumori,* 89**,** 586-93.

Pidsley, R., C. C. Y Wong, M. Volta, K. Lunnon, J. Mill & L. C. Schalkwyk (2013) A data-driven approach to preprocessing Illumina 450K methylation array data. *BMC Genomics,* 14**,** 293.

Raum, E., D. Rothenbacher, M. Löw, C. Stegmaier, H. Ziegler & H. Brenner (2007) Changes of cardiovascular risk factors and their implications in subsequent birth cohorts of older adults in Germany: a life course approach. *Eur J Cardiovasc Prev Rehabil,* 14**,** 809-14.

Riboli, E. (2001) The European Prospective Investigation into Cancer and Nutrition (EPIC): plans and progress. *J Nutr,* 131**,** 170S-175S.

Schwarzer, G., J. Carpenter & G. Rücker. 2015. *Meta-analysis with R*. Springer Publishing International.

Severi, G., M. C. Southey, D. R. English, C. H. Jung, A. Lonie, C. McLean, H. Tsimiklis, J. L. Hopper, G. G. Giles & L. Baglietto (2014) Epigenome-wide methylation in DNA from peripheral blood as a marker of risk for breast cancer. *Breast Cancer Res Treat,* 148**,** 665-73.

Smyth, G. K. 2005. limma: Linear Models for Microarray Data. In *Bioinformatics and Computational Biology Solutions Using R and Bioconductor,* eds. R. Gentleman, V. J. Carey, W. Huber, R. A. Irizarry & S. Dudoit, 397-420. New York, NY: Springer New York.

Steen, J., T. Loeys, B. Moerkerke & S. Vansteelandt (2017) Flexible Mediation Analysis With Multiple Mediators. *Am J Epidemiol,* 186**,** 184-193.

Stensrud, M. J. & M. A. Hernán (2020) Why Test for Proportional Hazards? *JAMA,* 323**,** 1401-1402.

Tchetgen, E. J. & I. Shpitser (2012) Semiparametric Theory for Causal Mediation Analysis: efficiency bounds, multiple robustness, and sensitivity analysis. *Ann Stat,* 40**,** 1816-1845.

Tchetgen Tchetgen, E. J. (2013) Inverse odds ratio-weighted estimation for causal mediation analysis. *Stat Med,* 32**,** 4567-80.

Teschendorff, A. E., F. Marabita, M. Lechner, T. Bartlett, J. Tegner, D. Gomez-Cabrero & S. Beck (2013) A beta-mixture quantile normalization method for correcting probe design bias in Illumina Infinium 450 k DNA methylation data. *Bioinformatics,* 29**,** 189-96.

Valeri, L., X. Lin & T. J. VanderWeele (2014) Mediation analysis when a continuous mediator is measured with error and the outcome follows a generalized linear model. *Stat Med,* 33**,** 4875-90.

VanderWeele, T. J. 2015. *Explanation in Causal Inference: Methods for Mediation and Interaction*. New York: Oxford University Press.

VanderWeele, T. J., L. Valeri & E. L. Ogburn (2012) The role of measurement error and misclassification in mediation analysis: mediation and measurement error. *Epidemiology,* 23, 561-4.

VanderWeele, T. J. & S. Vansteelandt (2014) Mediation Analysis with Multiple Mediators. *Epidemiol Methods,* 2, 95-115.

Whelan, B. J. & G. M. Savva (2013) Design and methodology of the Irish Longitudinal Study on Ageing. *J Am Geriatr Soc,* 61 Suppl 2, S265-8.

Wiksten, A., G. Rücker & G. Schwarzer (2016) Hartung-Knapp method is not always conservative compared with fixed-effect meta-analysis. *Stat Med,* 35, 2503-15.

Wilson, R., S. Wahl, L. Pfeiffer, C. K. Ward-Caviness, S. Kunze, A. Kretschmer, E. Reischl, A. Peters, C. Gieger & M. Waldenberger (2017) The dynamics of smoking-related disturbed methylation: a two time-point study of methylation change in smokers, non-smokers and former smokers. *BMC Genomics,* 18, 805.

Wong Doo, N., E. Makalic, J. E. Joo, C. M. Vajdic, D. F. Schmidt, E. M. Wong, C. H. Jung, G. Severi, D. J. Park, J. Chung, L. Baglietto, H. M. Prince, J. F. Seymour, C. Tam, J. L. Hopper, D. R. English, R. L. Milne, S. J. Harrison, M. C. Southey & G. G. Giles (2016) Global measures of peripheral blood-derived DNA methylation as a risk factor in the development of mature B-cell neoplasms. *Epigenomics,* 8, 55-66.

Zeilinger, S., B. Kühnel, N. Klopp, H. Baurecht, A. Kleinschmidt, C. Gieger, S. Weidinger, E. Lattka, J. Adamski, A. Peters, K. Strauch, M. Waldenberger & T. Illig (2013) Tobacco smoking leads to extensive genome-wide changes in DNA methylation. *PLoS One,* 8, e63812.

Zhang, Y., B. Schöttker, I. Florath, C. Stock, K. Butterbach, B. Holleczek, U. Mons & H. Brenner (2016) Smoking-Associated DNA Methylation Biomarkers and Their Predictive Value for All-Cause and Cardiovascular Mortality. *Environ Health Perspect,* 124, 67-74.

Zhang, Y., R. Wilson, J. Heiss, L. P. Breitling, K. U. Saum, B. Schöttker, B. Holleczek, M. Waldenberger, A. Peters & H. Brenner (2017) DNA methylation signatures in peripheral blood strongly predict all-cause mortality. *Nat Commun,* 8, 14617.
